# Optimization of Multi-Ancestry Polygenic Risk Score Disease Prediction Models

**DOI:** 10.1101/2024.04.17.24305723

**Authors:** Jon Lerga-Jaso, Andrew Terpolovsky, Biljana Novković, Alex Osama, Charlie Manson, Sandra Bohn, Adriano De Marino, Mark Kunitomi, Puya G. Yazdi

**Affiliations:** Research & Development, Omics Edge, Miami, FL, USA; Almaden Genomics, San Francisco, CA, USA

## Abstract

**Background:** Polygenic risk scores (PRS) have ushered in a new era in genetic epidemiology, offering insights into individual predispositions to a wide range of diseases. However, despite recent marked enhancements in their predictive power, there are still challenges that need to be overcome before PRS-based models can be broadly applied in the clinic, including sufficient accuracy, easy interpretability and portability across diverse populations.

**Methods:** Leveraging trans-ancestry genome-wide association study (GWAS) meta-analysis, we generated novel, diverse summary statistics for 30 medically-related traits which were used to benchmark the performance of six existing PRS algorithms using UK biobank. Observing that SBayesRC had the best overall performance but recognizing strengths in each method, we developed an ensemble PRS model using logistic regression to combine outputs from top-performing algorithms. This ensemble model was validated on the diverse eMERGE and PAGE MEC cohorts, and the performance was compared against current state-of-the-art PRS models. To enhance predictive accuracy for clinical application, we incorporated easily-accessible clinical characteristics such as age, gender, ancestry and risk factors, creating disease prediction models intended as prospective diagnostic tests, with easily interpretable positive or negative outcomes.

**Results:** Predictive performance of PRS models improved with trans-ancestry GWAS meta-analysis and was further enhanced by the ensemble model, which surpassed state-of-art PRS models. When applied to external cohorts, performance drops were minimal, indicating good calibration. After adding clinical characteristics, 12 out of 30 models surpassed 80% AUC. Further, 25 traits exceeded the diagnostic odds ratio (DOR) of 5 and 19 traits exceeded DOR of 10 for all ancestry groups, indicating high predictive value. The highest DOR in a population with a sufficient number of cases was 66.2 for Alzheimer’s disease in Europeans. Our PRS model for coronary artery disease identified 55-80 times more true coronary events than rare pathogenic variant models, reinforcing its clinical potential. The polygenic component modulated the effect of high-risk rare variants, stressing the need to consider all genetic components in clinical settings.

**Conclusions:** Newly developed PRS-based disease prediction models have sufficient accuracy and portability to warrant consideration of being used in the clinic.

## Background

Polygenic risk scores (PRSs) have emerged as a transformative tool in genetic epidemiology, harnessing the wealth of data generated by genome-wide association studies (GWAS) to predict an individual’s predisposition to complex diseases [1]. By aggregating numerous genetic variants, each contributing small to modest effects, PRS offers an understanding of genetic susceptibility across a spectrum of human diseases, including cardiovascular conditions, psychiatric disorders, and cancers [2-6]. The utility of PRSs extends beyond mere risk prediction; they hold promise for personalized medicine, where interventions can be tailored based on an individual’s genetic risk profile. Moreover, PRSs can enhance disease screening strategies, inform clinical decision-making, and potentially guide lifestyle and therapeutic interventions aimed at mitigating disease risk [1,3,7-9]. Despite this potential, the application of PRSs in clinical settings is in its nascent stages, grappling with challenges such as improving the accuracy and interpretability of scores, ensuring equitable performance across diverse populations, and integrating genetic risk information with environmental and lifestyle factors for a comprehensive approach to disease prediction and prevention [1,10,11].

Recent advancements in computational methods, alongside the exponential growth in GWAS sample sizes, have markedly enhanced the predictive power of PRSs [8,12]. However, developing and validating PRS models that are generalizable across different ancestries remains imperative, as current models often exhibit reduced efficacy in non-European populations [1,8]. Additionally, any PRS used in a clinical setting must demonstrate clinical utility and easy interpretability that will alter patient care decisions based on the results [13].

With those aims in mind, we set about to create and validate multi-ancestry PRSs that incorporate diverse clinical characteristics. We treated this task as a binary classification problem and systematically tested different combinations of inputs to make disease predictions that are easily interpretable and would alter physician intervention decisions. We build and evaluate different PRS models for 30 different medically-related traits starting from assessing the benefit of GWAS meta-analysis, to adding demographic and clinical risk factors to our models to maximize their predictive power.

## Methods

### Cohorts and other PRS models

**UK Biobank Cohort**: The UK Biobank used a custom Axiom genotyping array, which assayed 825,927 genetic variants for around 500,000 participants aged 40-69 years [14]. This was followed by comprehensive genome-wide imputation utilizing the Haplotype Reference Consortium (HRC) haplotype resource and the UK10K + 1KG reference panel, culminating in 96 million variants. From the imputed dataset, we isolated 13.7 million variants, selecting those with a minor allele frequency greater than 0.001 and a Hardy-Weinberg equilibrium *P* value exceeding 10^-10^. This carefully curated subset of high-quality variants served as the foundation for our PRS model development.

We first restrict individuals to the ones used for computing the principal components in the UK Biobank (Field 22020) and defined as the ‘White British ancestry’ group (UKB Data Field 22006). These individuals, called the White British Unrelated (WBU) subgroup, are a high quality set of unrelated individuals that underwent stringent quality controls including the exclusion of samples with an autosomal missing rate greater than 0.02, mismatches between inferred and self-reported sex, and outliers based on heterozygosity, as detailed in Bycroft et al. (2018) [14]. This subgroup was used by Thompson et al. (2024) [8] to generate GWAS summary statistics. We used the same subgroup (available at zenodo.org/records/6631952) as the UKB fraction that was then meta-analyzed with other GWAS datasets.

After excluding the previously identified WBU subset, we retrieved 104,604 remaining UKB samples for downstream analysis. We adopted this approach, which was proposed by Thompson et al (2024) [8], to increase the representation of non-European ancestries within this cohort for PRS training, testing and validation. We divided this cohort into a training set of 30,000 individuals and a testing set of 74,604 individuals for benchmarking the performance of each PRS algorithm. Subsequently, we split the testing set into two sub-cohorts: one consisting of 30,000 participants for retraining our novel ensemble PRS method and developing new risk models that integrate various PRSs and additional demographic factors, and another comprising 44,604 participants for testing these models. This structured approach enabled direct comparisons of our PRS results with those developed by Thompson et al., using the same subsets, thereby ensuring the comparability and robustness of our findings. All experiments and the cohorts and subsets used in this study are detailed in Fig. 1, which presents a schematic of the study design.

**Fig. 1.**
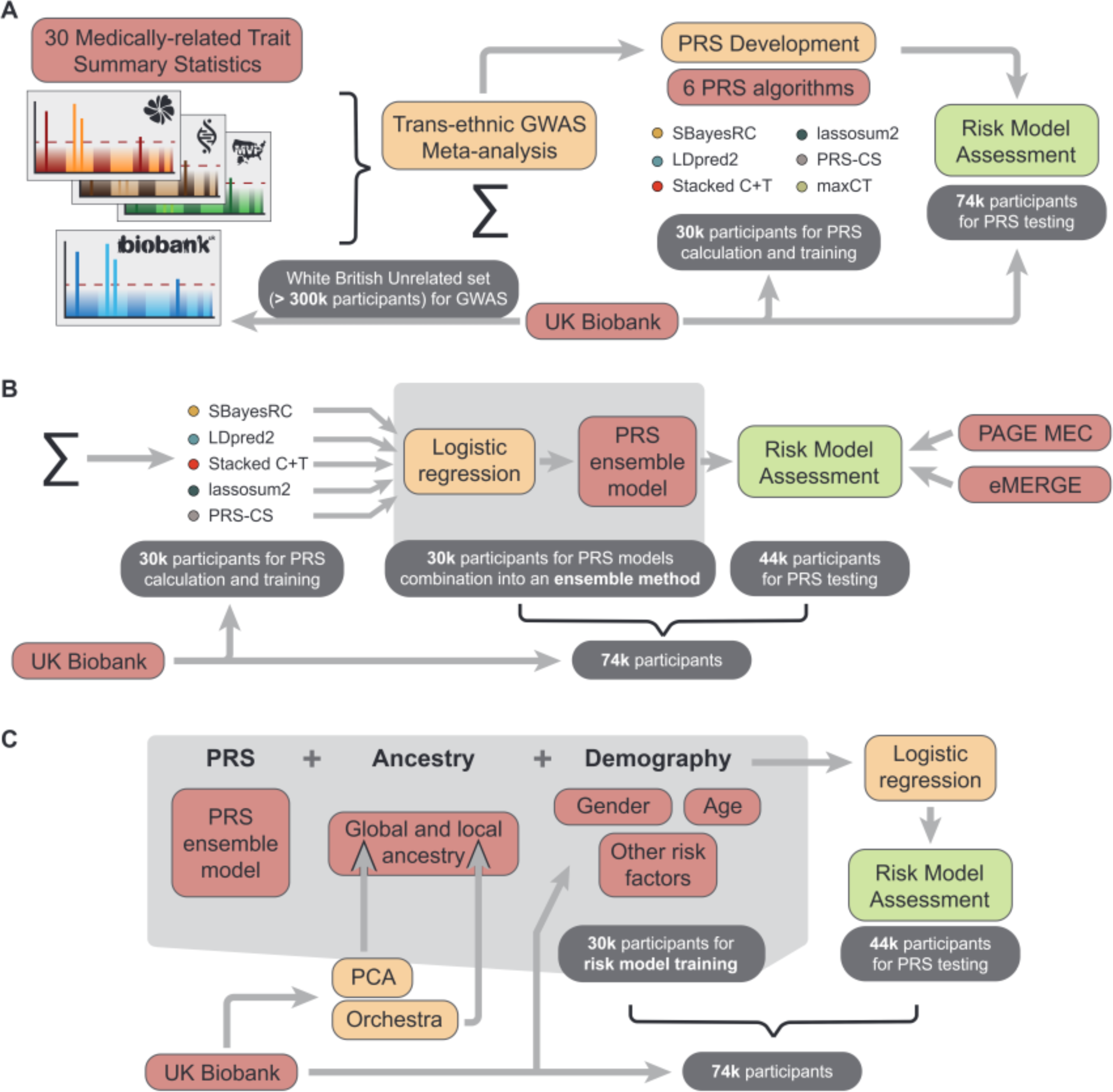
Study design schematics. **A)** Benchmarking study. Comparison of six PRS prediction algorithms applied to 30 medically-related traits. **B)** PRS evaluation study. Combination of 5 PRS models into an ensemble PRS model. maxCT was excluded since SCT is already a stacked model derived from maxCT PRS models. **C)** Final risk model assessment. We added additional information to enhance model risk prediction. See main text for details.

We also adopted the methodology outlined by Thompson et al (2024) [8] to infer genetic ancestry, ensuring our results were comparable. In this regard, data was standardized by initially grouping participants according to their self-reported ethnicity. In cases where multiple ethnicity entries were available, the primary entry was used unless it was missing, in which case secondary or tertiary entries were considered. Ethnicities were categorized as follows: African into “African”; Bangladeshi, Indian, and Pakistani into “South Asian”; Chinese into “East Asian”; other Asian backgrounds into “Asian”;various mixed and Caribbean backgrounds into “Mixed”;and British, Irish, and other white backgrounds into “European”. Categories such as “Prefer not to answer” were kept as “Unknown”. Centroid coordinates for “European”, “East Asian”, “South Asian”, and “African” ancestry groups were defined in the principal component (PC) space from the first four PC axes (see below for variant pruning used to derive principal components for risk models). We calculated cosine similarity between each individual’s PC score vector and each population centroid using the cosineSim function. Ancestry proportions were then estimated by applying a softmax transformation (base=exp(3)) to these cosine similarities, yielding a dataframe with one column per population centroid. Individuals were assigned a superpopulation category based on the highest ancestry proportion, ensuring no individual was labeled as admixed. Our results closely matched those reported by Thompson et al., with 9,501 individuals categorized as “African”, 2,931 as “East Asian”, 82,318 as “European”, and 9,854 as “South Asian”.

All UKB participants provided informed consent. Our research project (Project Application Number 84038) received approval from the UK Biobank.

**eMERGE Network Cohort**: Access to the eMERGE cohort was granted via dbGaP (phs001584.v2.p2), focusing on subsets c1, c4, c5, c6, c7, c8, and c10, which did not require Institutional Review Board approval [15]. This yielded data from 84,215 participants, featuring both genetic and phenotypic information, across diverse U.S. populations, including Blacks (or Africans; N = 10,126), Asians (N = 920), Whites (N = 67,316), Latinos (N = 3,303), Native Americans (N = 101) and Pacific Islanders (N = 6). Individuals with unknown self-reported ethnicity were excluded. Due to relatively small numbers, Native Americans were grouped with Latinos, and Pacific Islanders with Asians. Despite the genetic dataset containing over 39 million variants, it lacked certain SNPs from our models and those developed by Lennon et al. (2024) [16] (see below). To address this issue, we phased the newly released whole genome sequencing data from the UKB (UKB Dragen WGS), available on the UK Biobank Research Analysis platform, with ShapeIt5 [17] and used it as our reference panel. This enabled us to impute missing SNPs, ensuring minimal SNP loss when applying various PRS models. The eMERGE cohort was lifted over to GRCh38 with Picard tools [18] and then normalized with BCFtools [19]. Imputation was performed using Beagle5.4 [20] with default parameters.

**PAGE MEC Cohort**: Access to the PAGE MEC [21] cohort was granted via dbGaP (phs000220.v2.p2). We prioritized individuals genotyped using the MEGA Consortium array over the Metabochip due to its significantly larger number of variants (1,705,969 vs. 196,725 variants). This choice was made to guarantee more effective imputation and enhance the reliability of our analyses. In total, this yielded data from 9,098 participants, featuring both genetic and phenotypic information, across diverse U.S. populations, including Blacks (or Africans; N = 3,520), Hawaiians (N = 2,104), Japanese (N = 3,451) and Latinos (N = 23). Individuals with unspecified self-reported ethnicity were excluded from the study, along with Latinos, who were omitted due to their relatively small numbers. Similar to the eMERGE cohort, we imputed the PAGE MEC cohort to reduce SNP loss, ensuring the effective application of various PRS models. Finally, Hawaiian and Japanese individuals were combined into an Asian group when necessary to facilitate performance comparisons with groups from other cohorts.

**Other PRS models:** We compared the PRS models we developed to those of Thompson et al (2024) [8], who conducted a systematic evaluation of their scores against numerous previously published models. Additionally, we compared our models to those of Lennon et al. (2024) [16], who endeavored to validate high-quality models for 10 diseases. These comparisons are particularly pertinent in the field of PRS due to the demonstrated enhanced power of these models, which suggests their medical actionability and potential clinical utility. Thompson et al.’s PRS profiles in UKB were retrieved from the UK Biobank Research Analysis platform, while Lennon et al.’s PRS models can be accessed from here: github.com/broadinstitute/eMERGE-implemented-PRS-models-Lennon-et-al. Thompson et al. provided PRSs exclusively for UKB samples without accompanying models, limiting our comparative analysis to the UKB dataset. In contrast, Lennon et al. provided PRS models for 10 diseases, eight of which overlapped with ours, applicable to cohorts other than the UKB, which was used for model training. Consequently, we applied these models to the eMERGE cohort, where they were initially tested. The use of the UKB whole genome sequencing reference panel for eMERGE imputation ensured minimal SNP loss (< 1%) when implementing both our and Lennon et al.’s PRS models, even when liftover was necessary.

### Selection of Phenotypes and Definitions

For our study, we adopted the 28 diseases listed in Thompson et al (2024) [8], leveraging the extensive and richly phenotyped cohorts available for these conditions. We excluded multiple sclerosis due to a lack of additional summary statistics that showed improvement over random predictions (AUC not significantly above 50%), resulting in a final list of 27 diseases. Additionally, we incorporated three quantitative health-related traits —body mass index, total cholesterol and triglyceride levels—which were transformed into binary traits for consistent analysis. We defined hypercholesterolemia and hypertriglyceridemia as values above 200 mg/dL, and obesity as a body mass index above 27.5 for Asian individuals and above 30 for all other ethnicities. This adjustment expanded our focus to 30 clinical traits.

Trait definitions were aligned with those presented in Thompson et al. (2024) [8], where phenotype variables for each trait in the UK Biobank were derived from a combination of self-reported responses and ICD-10 records, detailed in Table S1. For type 1 and 2 diabetes, we employed a more detailed case definition process. Initially, we designated type 1 diabetes cases using the UK Biobank code 1222 in field 20002 and the ICD-10 code E10, and type 2 diabetes using UKB code 1223 and ICD-10 code E11, as per Privé et al. (2019) [22]. We excluded other diabetes forms, setting them as missing using codes 1220-1223 and ICD-10 codes E10-E14. Subsequently, we implemented the decision tree methodology outlined in Thompson et al. (2024) to refine these classifications: (i) Individuals without any diabetes-related labels, self-reports, ICD-10 codes, or diabetes medications (including insulin product, metformin, rosiglitazone and metformin, troglitazone, pioglitazone, rosiglitazone, glimepiride, glibenclamide, glibornuride, gliclazide, glipizide product, glipizide, gliquidone, chlorpropamide, acetohexamide, tolazamide, tolbutamide, diabinese, minodiab, diabetamide, repaglinide or nateglinide) were categorized as controls; (ii) Female participants who reported only gestational diabetes, without other diabetes types or medications, were excluded from the analysis; (iii) Participants reporting diabetes and on non-insulin, non-metformin diabetes medications, or diagnosed after age 36, were classified as having type 2 diabetes; (iv) Those reporting a type 1 diabetes diagnosis or using insulin were assigned a type 1 diabetes status; (v) All other individuals with a diabetes label but not fitting the above categories were assigned type 2 diabetes status. Next, we reassessed the initial definitions from the UK Biobank code in 2002 and ICD-10 codes, and the status obtained with the decision tree approach, adjusting for discrepancies: (i) Where control and diabetic case classifications conflicted, individuals were designated as missing due to reliability concerns; (ii) Type 1 diabetes classification were recovered from previously missing values only if there was an ICD-10 code for individuals diagnosed before 25 years of age and who were on insulin only; for type 2 diabetes, recovery of cases from missing data occurred for those with an unspecified type but diagnosed after age 50, or if the date of type 1 diabetes diagnosis was at least a year later to the type 2 diagnosis, or they were taking non-metformin oral diabetes medications. Finally, in PRS analyses for diabetes, we excluded individuals diagnosed with the alternate diabetes type to ensure a clear and accurate set of cases and controls without confounding factors.

We utilized both ICD-9 and ICD-10 codes in the eMERGE dataset to define cases and controls. The presence of these codes categorized participants as cases for specific diseases. Additionally, the cohort data already included identified cases for conditions like glaucoma, asthma, age-related macular degeneration, venous thromboembolism, and type 2 diabetes, which facilitated the identification of further cases. Individuals on statins following a Major Adverse Cardiovascular Event were excluded from the hypercholesterolemia and hypertriglyceridemia analyses, and those diagnosed with type 2 diabetes were removed from type 1 diabetes analyses, and vice versa. Following the methodology of Lennon et al. (2024) [16], we restricted case definitions to certain age ranges that reflect typical disease onset. Specifically, diagnoses of type 1 diabetes, asthma and Crohn’s disease were considered positive if made between ages 3 and 17; obesity, type 2 diabetes, celiac disease, psoriasis, schizophrenia and bipolar disorder were diagnosed across ages 3 to 75; other diseases were categorized as late-onset (over 17 years old). Individuals outside the age range of 3 to 75 were excluded.

In the PAGE MEC cohort, baseline reports of diabetes, high blood pressure, heart attack, stroke, body mass index, total triglycerides, and total cholesterol were respectively categorized as type 2 diabetes (DIABET variable), hypertension (HIBP), coronary artery disease (HATTACK), ischemic stroke (STROKE), obesity (BMI), hypertriglyceridemia (_triglyc) and hypercholesterolemia (_tot_chol). We refined our identification of additional type 2 diabetes cases by using the “med_diab” variable, which reported type 2 diabetes medications taken at the time of blood draw. Cardiovascular disease was also defined by the presence of both heart attack and stroke. The analysis excluded total triglycerides due to a low sample size. Analyses involving epithelial ovarian cancer and breast cancer were conducted exclusively with female participants, while studies on prostate cancer were restricted to male individuals.

### GWAS Meta-Analysis

We generated new genome-wide summary statistics for the 30 health-related conditions covered in this article, utilizing a trans-ancestry GWAS meta-analysis approach that incorporated data from various cohorts, including the UK Biobank, the Finngen and the BioBank Japan, among others. Detailed information about all the studies collected is provided in Table S2 [14,23-73].

We utilized the METAL[74] software to conduct GWAS meta-analyses employing two specific schemes. For effect size estimates (beta coefficients) and standard errors, we adopted the SCHEME STDERR, which processes effect size estimates and standard errors from each study to compute aggregate effects of each variant. Ensuring uniform units for effect sizes across all included summary statistics was essential for consistency. For *P* value computation, we implemented the default SCHEME SAMPLESIZE, which adjusts *P* values and directional effects based on sample size. In this context, we calculated the effective GWAS sample size for binary traits using the formula 4 / (1 / Ncases + 1 / Ncontrols) where Ncases and Ncontrols represent the number of cases and controls in the GWAS, respectively. For quantitative traits, we directly used the total number of samples. This dual-mode approach provided flexibility in our analysis, as the SAMPLESIZE model is capable of estimating corrections for sample overlap, thereby accommodating such corrections in meta-analyses where overlaps are inevitable. Consequently, we sourced our *P* values from ‘SAMPLESIZE metaanalysis’ and beta coefficients from ‘STDERR metaanalysis’, effectively addressing the complexities of our data.

We sourced summary statistics from publicly available GWAS data, with the GWAS Catalog [75] serving as a predominant source. Additionally, we accessed data from specific project platforms such as the Finngen, BioBank Japan, Global Lipids Genetics Consortium and others designed for specific studies. Access was also requested from dbGaP to obtain a dataset of summary statistics for melanoma (phs001868.v1.p1 [48]), as well as data from the Million Veterans Program (phs001672.v11.p1 [66]). To minimize overfitting, we excluded studies incorporating the UKB or used versions of summary statistics that deliberately excluded UKB samples, even if the broader study included them in the corresponding article. From the several genetic models presented by Guindo-Martínez *et al*. (2021) [27], we selected the additive model for our analyses.

The earliest dataset used was from Lambert et al. (2013) [29] for Alzheimer’s disease, with subsequent datasets being more recent, from 2015 onward. We excluded studies if they presented fewer than 100,000 variants or were confined to a single chromosome or region. Moreover, older studies were removed if they were included in more recent published releases or meta-analyses, used in our study. We also conducted correlation analyses between studies on nominally significant variants to ensure reliable comparison of effects to verify beta direction consistency. Instances of complete inverse or totally non-correlative beta directions between UKB and other studies indicated potential labeling errors for effect alleles or allele assignments. We noticed that significantly associated SNPs with allele frequencies close to 50% often displayed such inconsistent beta directions in studies that did not correlate with others, implying potential mislabeling of minor alleles as effect alleles in the GWAS data. We harmonized the summary statistics by annotating SNP IDs from dbSNP version 155 and calculated betas from logarithms of odds ratios where this was the sole metric reported. Standard errors were derived by dividing the absolute beta coefficients by their corresponding z-scores, which were computed from *P* values using the inverse of the standard normal cumulative distribution. If a *P* value was below the threshold of 10^-308^, we set it to this value as it represents the lower limit that METAL can process as input. Additionally, we performed liftover processes from GRCh38 to GRCh37 genome build and implemented other necessary harmonization steps to ensure consistency across our datasets.

### PRS Model Benchmarking

To benchmark predictive performance and elucidate each method’s strengths and weaknesses, we compared the following PRS algorithms: PRS-CS [76], SbayesRC [77], PolyPred [78], NPS (partitioning-based non-parametric shrinkage) [79], lassosum2 [80], LDpred2 [81], maxCT and Stacked C+T (SCT) [22]. The training set was used to determine the optimal hyper-parameters for PRS-CS, NPS, PolyPred, lassosum2, LDpred2, and maxCT, in addition to the stacking weights for SCT. SBayesRC was trained using only summary statistics, eliminating the need for individual-level data.

To implement PolyPred, we integrated two complementary predictors—PolyFun-pred and SBayesRC outputs—and applied this model to five traits: asthma, age-related macular degeneration, ulcerative colitis, venous thromboembolism, and cardiovascular disease. However, the performance gains from PolyPred were minimal, improving by only about 0.1% over the results obtained from SBayesRC alone. Additionally, PolyPred was computationally demanding, being roughly 75 times more resource-intensive than SBayesRC. This increase in computational load is likely due to its extensive linkage disequilibrium (LD) reference data requirements, where SBayesRC utilizes a 50 GB LD panel, in contrast to PolyPred’s 2.9 TB LD matrices. We also evaluated the NPS software for the same set of traits and encountered similar challenges, including the need for over 3 TB of local storage for dosage matrices, prolonged processing times, and comparatively lower accuracy among the PRS methods we reviewed. The artificially high AUC observed during training is typically indicative of model overfitting. Given these findings, coupled with faster and more accurate performance from other methods, we decided against including PolyPred and NPS in our final benchmarking analysis.

For maxCT, tuning parameters included a squared correlation (*r²*) threshold for clumping and a base size for the clumping window, with default settings specified in Privé et al. (2019) [22]. Additionally, a series of 50 thresholds on *P* values were applied, ranging from the least to the most significant in the final meta-analyzed summary statistics, and evenly distributed on a log-log scale. LDpred2’s tuning involved selecting the proportion of causal SNPs from a sequence of 21 logarithmically spaced values ranging from 10^-5^ to 1, and inferring the per-SNP heritability chosen from {0.3, 0.7, 1, 1.4} times the total heritability, as estimated by LD score regression. We enabled the ‘sparse’ option to truncate minor effects to zero, which effectively doubled the number of models considered for training by incorporating both sparse and non-sparse models. lassosum2’s tuning parameters included the lasso penalty lambda (L1-regularization), selected from 30 values logarithmically spaced between lambda0 -the maximum value where all coefficients are zero, derived from the maximum absolute standardized beta coefficient- and 1% of lambda0, along with a delta (L2-regularization) parameter for the LD matrix chosen from {0.001,0.01,0.1,1}. We developed the PRS-CS PRS model using default settings for the gamma-gamma prior parameters (a = 1 and b = 0.5) calculated on a per-chromosome basis. We did not specify the global shrinkage parameter phi, allowing PRS-CS to learn phi from the data through a fully Bayesian approach, which is advisable in the context of large GWAS sample sizes such as ours [75]. SBayesRC was executed using its default settings, enabling the software to determine the optimal estimates for heritability and the count of non-zero effect variants. Additionally, we adjusted the eigen variance cutoffs for model tuning in response to software-generated warnings. The original threshold set of (0.995, 0.99, 0.95, 0.9) was expanded to (0.995, 0.9, 0.8, 0.7, 0.6) to accommodate findings that suggested the optimal tuning parameter was near the minimum threshold, thus necessitating lower cutoff values.

Both LDpred2 and lassosum2 were implemented using the functions *snp_ldpred2_grid* and *snp_lassosum2* from the R package *bigsnpr*, respectively. We adopted an LD radius of 3 cM to approximate local LD patterns, which assumes that variants further away than this distance are not correlated. Moreover, genetic markers were further restricted to the HapMap3 panel, following recommendations by Privé et al. (2021) [81]. Conversely, maxCT and SCT were executed using corresponding functions from the same R package (*snp_grid_clumping* and *snp_grid_PRS*, and *snp_grid_stacking*, respectively). However, for these analyses, we limited the set of genetic variants to those achieving a significance level of *P* < 0.1 in the meta-analysis. PRS-CS scores were also based on HapMap3 sites, as precomputed by the PRS-CS authors. In this case, we opted for data from the UKB as our reference panel for LD after it showed improved results in preliminary testing across several traits compared to those using the 1000 Genomes Project. PRS-CS is a Python based command line tool (available here: github.com/getian107/PRScs) and final PRSs were computed from the SNP weights output using the PLINK --score command. We utilized SBayesRC (available at github.com/zhilizheng/SBayesRC), applying the LD reference supplied by the authors from the UKB. For our analysis, we employed two different sets of SNPs: the HapMap3 set and a larger set consisting of 7 million SNPs, both incorporating functional genomic annotations to enhance polygenic prediction. During an initial testing phase across various traits, the 7 million SNP panel demonstrated superior performance compared to the HapMap3 set. Thus, we selected the 7 million SNP set for benchmarking and further analyses.

The models with the highest prediction accuracy on the training set were selected based on the Area under the ROC curve (AUC). The predictive performance of these final models was then evaluated on the independent testing set. Unlike diseases encoded as binary outcomes (case/control), optimal values for the quantitative traits – body mass index, total cholesterol, and triglyceride levels – were initially determined by training models to maximize the predictive *R²* between observed and predicted traits. Subsequently, these traits were converted to binary outcomes to facilitate AUC calculation and comparison with the testing samples. We sample 10,000 bootstrap replicates of the individuals in the testing set and compute the AUC for each of these. We then report the mean of these 10,000 values, along with their quantile at 2.5% and at 97.5% to act as the 95% confidence interval (CI) for the AUC. This is implemented in the function *AUCBoot* of R package *bigstatsr*. Additional metrics we employed include odds ratios that compare the top percentage of a population against the remainder, and odds ratios per standard deviation of the PRS distribution. For the latter, given that logistic regression aims to distinguish between cases and controls by outputting a probability rather than a risk distribution, we applied a rank-based inverse normal transformation to our ensemble PRS model. This approach was taken to ensure that the PRS distribution adheres to a normal distribution while maintaining the rank order of the data points.

### Multi-algorithm ensemble PRS method

We developed new PRS models for each disease by integrating scores previously obtained from SBayesRC, PRS-CS, SCT, LDpred2, and lassosum2. These models were retrained using logistic regression, employing the second training set to optimize the integration of these scores. This approach allowed us to harness the strengths of each individual scoring method to enhance overall predictive accuracy. Furthermore, this strategy proved to be an effective method of leveraging a larger cohort for training without the need to directly retrain the more computationally intensive and time-consuming PRS algorithms described above, which had been previously calibrated within a smaller, yet adequate cohort, thereby streamlining the overall process.

We optimized the logistic regression models using the *glmnet* method for binary classification, configured with 5-fold cross-validation to enhance reliability. The models were implemented in R using the *caret* package. Model training was controlled through the *trainControl* function, set to compute class probabilities and evaluate model performance based on the AUC metric. Hyperparameters were finely tuned using a grid search across a range of values for alpha (0 to 1, in 11 steps) and lambda (10^−4^ to 10^1^, in 50 logarithmic steps), which regulate the balance between L1 and L2 regularization. This approach allowed us to systematically explore the parameter space to maximize the AUC metric, ensuring optimal discrimination of binary (case/control) outcomes.

### The Addition of Ancestry Information and Clinical Characteristics to PRS-Based Disease Risk Models

To improve the predictive accuracy of our PRS models, we incorporated ancestry information through an iterative process. We began by incorporating the first four principal components (PCs) identified from a principal component analysis (PCA) on variants that were pruned to ensure linkage equilibrium. First, we removed strand-ambiguous SNPs A/T and G/C, then filtered the SNP dataset to exclude those with a minor allele frequency (MAF) below 2%, those failing the Hardy-Weinberg equilibrium test with a *P* value below 1×10^−6^ and those with over 5% missing data. After these quality control steps, we applied genotype pruning using a window size of 1000 kb, a step size of 50 SNPs, and an *r^2^* threshold of 0.2, which allowed us to capture broad ancestry patterns through PCA. We then enhanced these models by integrating ancestry estimates from Orchestra [82], a method for local ancestry deconvolution. We aggregated the results from all genomics regions into vectors that quantify the percentage of each ancestry assessed for every individual. Consequently, each individual’s genetic makeup was represented by a vector of values, with each vector element indicating the proportion of a specific ancestry. This approach allowed us to incorporate a series of columns into our model – each corresponding to a different ancestry – showing the percentage composition of that ancestry for each participant and enabling precise genetic profiling per individual. Orchestra’s results were provided at level 2, which offers a regional granularity intermediate between population-specific and continental scales of ancestry detail. Both measures were incorporated into a logistic regression model and trained in the second training set.

We also incorporated age, sex, and other clinical characteristics, easily gatherable in a clinical setting (Table S3). Age was calculated as the age at the earliest recorded diagnosis, using the earliest diagnosis date from the various sources outlined in Table S1, in conjunction with the date of recruitment and the age at recruitment. We chose characteristics that are known risk factors for each medically-related trait. We added this information to PRS and ancestry to make a final logistic regression model that would serve as a prospective diagnostic test, with an easily interpretable positive or negative value, typical of other diagnostic tests used in a clinical setting.

We employed the same approach as described previously for the logistic regression model, using grid-search techniques to fine-tune the regularization parameters, thereby ensuring the model’s generalizability.

### PRS profiles in clinical settings

We utilized whole exome sequencing data from the UK Biobank to analyze genetic risks for breast cancer, bowel cancer and coronary artery disease by identifying significant mutations in genes associated with these conditions. To identify carriers, we aggregated mutations across all pertinent genes for each disease type. Specifically, we used a list from Fahed et al. (2020) to pinpoint mutations in genes associated with familial hypercholesterolemia (*APOB*, *LDLR*, *PCSK9*), breast cancer (*BRCA1*, *BRCA2*), and colorectal cancer (Lynch syndrome genes: *MSH2*, *MSH6*, *MLH1*, *PMS2*) [2]. We expanded the list of genetic variants by including additional mutations identified as likely pathogenic or pathogenic in the ClinVar database. For familial hypercholesterolemia (FH) genes, we incorporated variants from Chora et al. (2018) [83], which adhered to diagnostic guidelines set by the American College of Medical Genetics and Genomics and the Association for Molecular Pathology [84]. We also added more variants for the *APOB* gene from ClinVar that are associated with hypercholesterolemia (search: apob[gene] and Hypercholesterolemia). For breast cancer, our list was enriched with mutations from the *ATM* and *PALB2* genes, classified as likely pathogenic and pathogenic in ClinVar under the category of familial breast cancer (X[gene] and familial cancer of breast). Additionally, we included the *CHEK2* 1100delC mutation, known for its significant association with breast cancer risk.

To evaluate the risk levels of individuals at the highest percentiles of their PRS profiles compared to carriers of pathogenic mutations, we focused our analysis on European individuals. This approach allowed us to use cumulative incidence plots for carriers within the full UKB cohort, maximizing sample size and statistical power due to the low frequency of these mutations, and minimizing confounding factors related to ancestry differences between the overall cohort and our UKB testing group, where PRS distribution percentiles were calculated. We determined the PRS percentile that matched the risk associated with having a pathogenic mutation, indicating that individuals with a PRS above this threshold have a comparable overall risk level to mutation carriers. To ascertain the accuracy of our findings, we calculated confidence intervals using a binomial test.

## Results

### Trans-ethnic GWAS Meta-analysis and PRS Algorithm Benchmarking

We generated novel genome-wide summary statistics for 30 medically-related traits, leveraging trans-ancestry genome-wide association study (GWAS) meta-analyses of diverse cohort data, including the UK Biobank (Fig. 1A). The following traits were selected: age-related macular degeneration, Alzheimer’s disease, asthma, atrial fibrillation, bipolar disorder, body mass index (BMI), bowel cancer, breast cancer, cardiovascular disease, celiac disease, coronary artery disease, Crohn’s disease, epithelial ovarian cancer, hypertension, ischemic stroke, melanoma, osteoporosis, Parkinson’s disease, primary open angle glaucoma, prostate cancer, psoriasis, rheumatoid arthritis, schizophrenia, systemic lupus erythematosus, total cholesterol, triglycerides, type 1 diabetes, type 2 diabetes, ulcerative colitis, venous thromboembolic disease. For the purpose of PRS evaluation analogous to diseases, the continuous traits – BMI, total cholesterol and triglycerides – were converted into binary outcomes: obesity, hypercholesterolemia, and hypertriglyceridemia, respectively.

The sample sizes and contributions of different studies to our meta-analyses are summarized in Fig. S1 and Table S2. For binary traits, the median number of cases and controls across all studies is 42,768 and 821,539, respectively. For quantitative traits, the median sample size is 1,205,118 individuals. In the meta-analyses, the number of cases for binary traits is 6.4 times greater than that of the UK Biobank (UKB) alone, while the number of controls is 3.1 times greater. For quantitative traits, the meta-analyses sample size is 7 times larger compared to using only the UKB summary statistics. Key contributors to the meta-analyses include the UKB, which accounts for 15.73% of cases and 32.1% of controls for binary traits, and 14.4% of samples for quantitative traits; the FinnGen project, which provides 26.5% of cases and 30.2% of controls for binary traits, and 11.8% of samples for quantitative traits; and studies involving the BioBank Japan, which contribute 4.9% of cases, 8.9% of controls for binary traits, and 4.7% of samples for quantitative traits. Notably, the Global Lipids Genetics Consortium is a major contributor for quantitative traits, accounting for 67.8% of the samples, due to the inclusion of total cholesterol and triglycerides levels in the analysis.

GWAS meta-analyses were conducted using METAL [74], incorporating publicly available external GWAS sources, predominantly sourced from the GWAS Catalog [75]. We kept out a set of 104,604 individuals from the UKB to serve as a training and testing cohort for PRS prediction and downstream validations (see Fig. 1A). The cohort was composed of 30,000 in the training and 74,604 in the testing set. We adopted the approach described by Thompson et al (2024) [8], which aimed to maximize the representation of non-European ancestries in this UKB subset of samples. The remainder of the samples, classified as the White British Unrelated (WBU) subset in the UKB project [14], were utilized for the GWAS component of the meta-analyses. This strategy facilitated direct comparisons of our results against the PRSs developed by Thompson et al. using the same testing set (see Methods section for more details).

To benchmark predictive performance and elucidate each method’s strengths and weaknesses, we trained six PRS algorithms: lassosum2 [80], LDpred2 [81], PRS-CS [76], SbayesRC [77], maxCT and Stacked C+T (SCT) [22]. Although we initially also trained PolyPred [78] and NPS (partitioning-based non-parametric shrinkage) [79], these were excluded from the final benchmarking because their performance did not enhance the results compared to the other methods, in addition to having longer computational training times (data not shown). SBayesRC was evaluated using both a subset of SNPs from the HapMap3 panel and an expanded set of over 7 million common SNPs. The latter yielded superior results and was selected for the benchmarking. PRS performance was evaluated using the area under the receiver operating characteristic curve (AUC), with the best results for each trait ranging from 83.4% (type 1 diabetes) to 55.9% (epithelial ovarian cancer), and a median of 68.1% (Fig. 2A). SBayesRC outperformed the other models for 21 out of 30 medical traits, 11 of which were statistically significant. In contrast, SCT and LDpred2 were superior in three traits each, with two of the SCT traits reaching statistical significance. Lassosum2 and PRS-CS each excelled in two and one trait, respectively (Fig. 2A and B). Moreover, when we relaxed our criterion to identify phenotypes where two models significantly outperformed the other four PRS methods (rather than just one model surpassing all others), we identified seven additional traits. In six of these cases, SBayesRC was one of the two top-performing models.

**Fig. 2.**
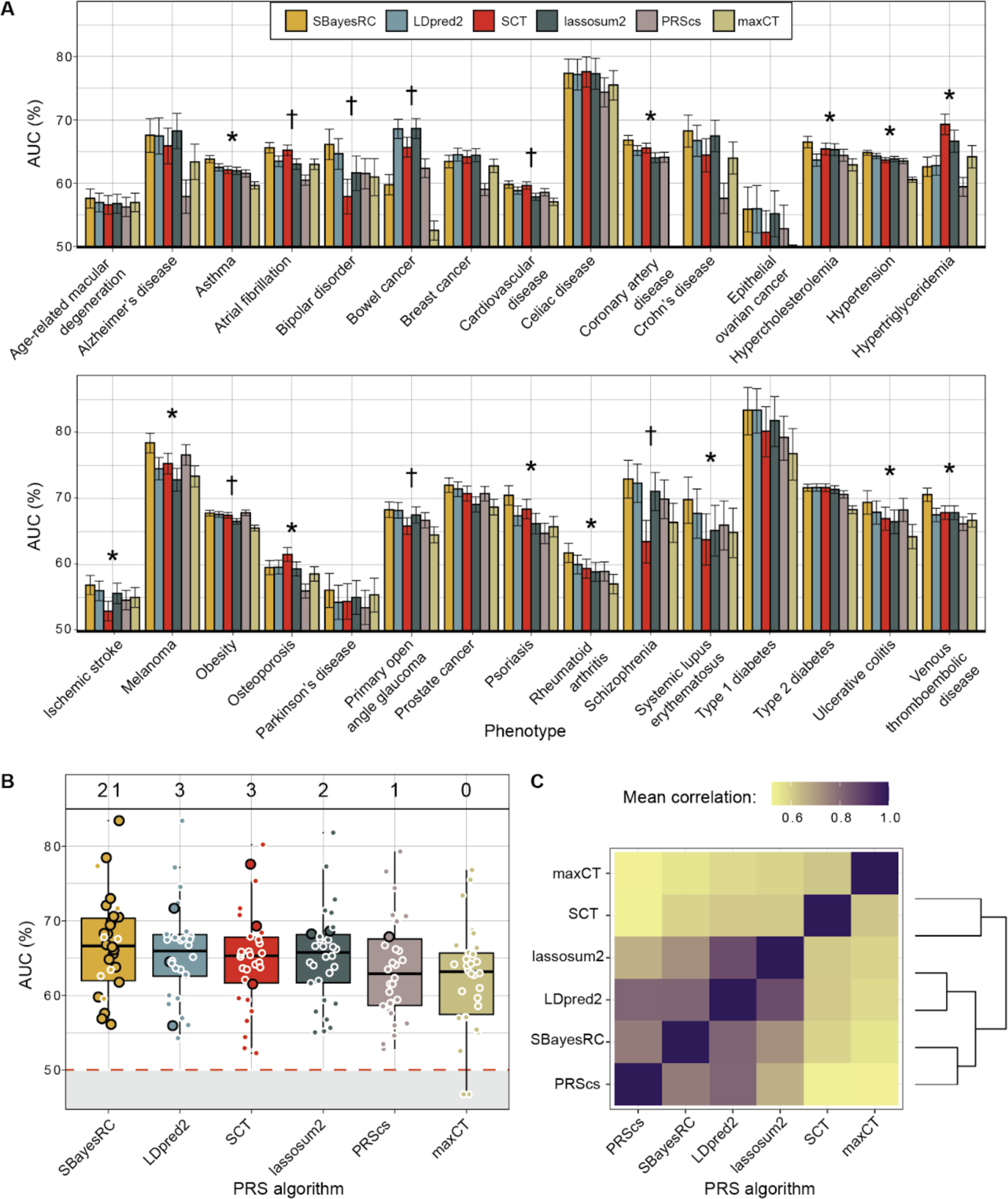
Benchmarking of six PRS prediction algorithms applied to 30 medically-related traits. **A)** Prediction accuracy using area under the curve (AUC) as a performance metric in the testing set of UKB. Conditions marked with an asterisk indicate those where SBayesRC or SCT significantly outperformed the other PRS algorithms. Conditions identified with a dagger represent a relaxed criterion where two algorithms significantly exceed the performance of the others. Error bars present the mean and 95% confidence interval of 10,000 non-parametric bootstrap replicates. DeLong’s test was used to compare the significant difference between AUC results across methods. Performance for breast and prostate cancer was calculated using only female and male individuals, respectively. **B)** Prediction results summarized per algorithm (with each dot representing one trait). The numbers at the top of the plot denote the count of traits where each PRS method demonstrated optimal performance, as indicated by highlighted dots. **C)** Mean pearson correlation coefficient (*r*) for PRS across the 30 traits between pairs of algorithms, along with a dendrogram depicting hierarchical clustering.

Pairwise correlations of PRS generated by each algorithm across all traits revealed high correlations between lassosum2 and LDpred2, as well as between LDpred2 and the other two Bayesian regression-based PRS algorithms (PRS-CS and SBayesRC), whereas correlations with SCT were less pronounced (Fig. 2C). We hypothesized that integrating insights from different models would lead to enhanced predictive accuracy.

### Predictive Performance of PRS in UK Biobank, eMERGE and PAGE MEC Cohorts

Building on the idea of integrating outputs from multiple PRS algorithms, we retrained a PRS model using logistic regression incorporating the scores previously obtained from SBayesRC, SCT, LDpred2, PRS-CS and lassosum2. We did not include maxCT since SCT is already a stacked version derived from different clumping plus thresholding models. To accomplish this, we partitioned the previously used UKB testing cohort into two subsets, creating an additional set of 30,000 participants for retraining the combination of PRS while retaining a separate group for testing the newly constructed models (44,604 participants) (see Fig. 1B). We assessed the performance of the newly constructed PRS models by comparing them to SBayesRC, which was identified as the top-performing individual PRS algorithm in our prior evaluations. Additionally, we contrasted these results derived from meta-analyzed summary statistics with those obtained solely using the UKB WBU GWAS component for PRS training with SBayesRC. These findings were benchmarked against the PRS reported by Thompson et al (2024) [8], who demonstrated that their PRS release surpassed a comprehensive array of previously published PRS models, and Lennon et al. (2024) [16], where the authors focused on optimizing and validating disease PRS models for clinical use.

Our ensemble model, which combined the outcomes from five PRS algorithms, significantly outperformed SBayesRC in our UKB testing cohort, leveraging both UKB-only and meta-analysis summary statistics (Fig. 3A and C). This combined method exceeded the SBayesRC algorithm for 23 out of 30 traits (*P* = 0.005, sign test). Moreover, our results proved superior to those reported by Thompson et al. [8] for 26 traits (*P* = 6 x 10^-5^, sign test), demonstrating significant improvements in AUC for 22 of these traits (DeLong’s test for AUC, *P* < 0.05), where our median AUC was 5.07% higher. The only exception was Parkinson’s disease, where Thompson et al. achieved significantly better results, albeit with a minor difference (*P* = 0.03, DeLong’s test). PRS analyzes utilizing only summary statistics from the UKB WBU set yielded the poorest performance relative to other strategies (median AUC 62.9% with UKB-only vs 66.9% with meta-analysis and 68.8% with ensemble method). However, results exclusively based on the UKB data showed marginally better outcomes for celiac disease. These negligible improvements may be attributable to random variation within the analysis.

**Fig. 3.**
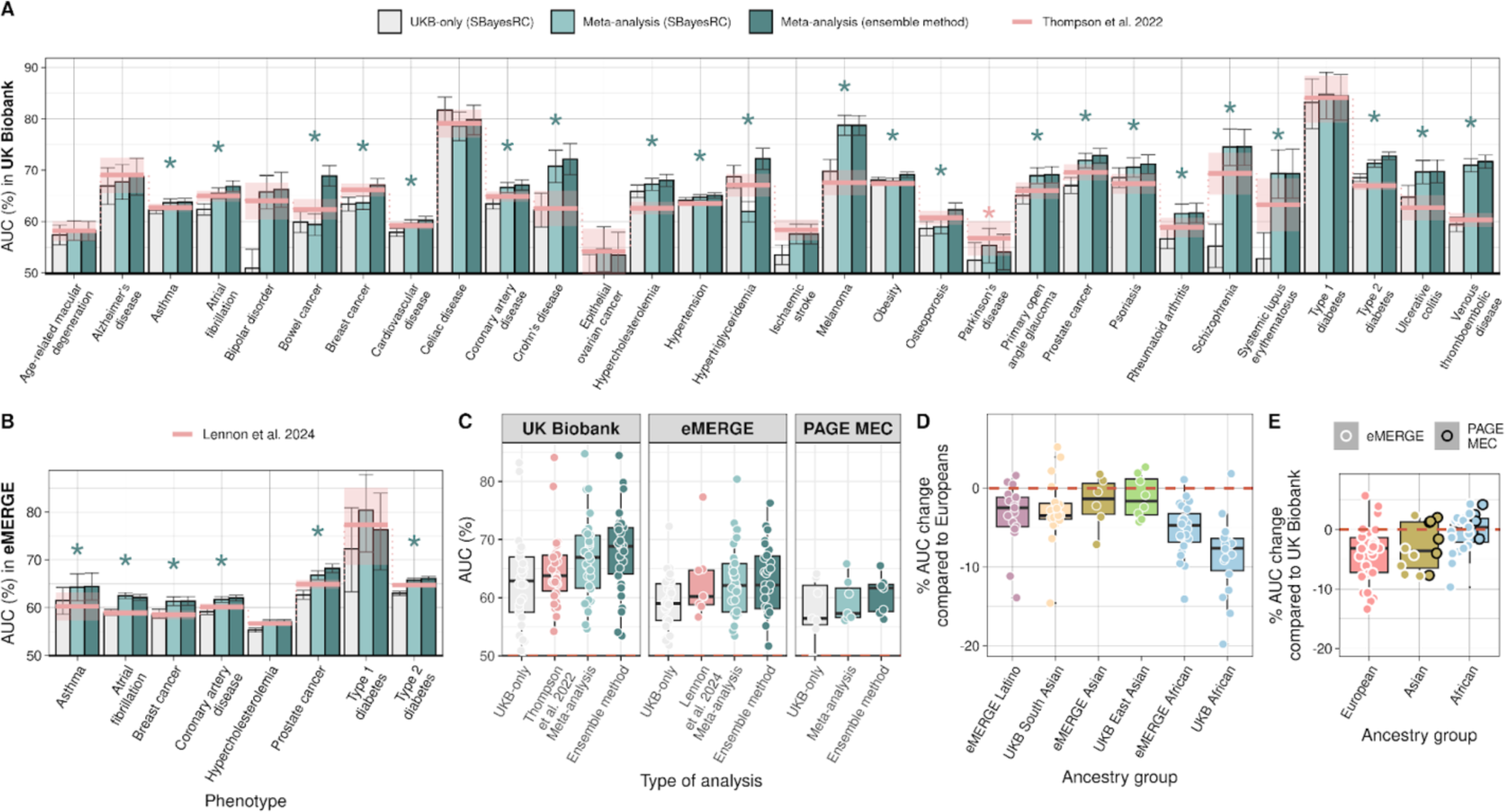
Predictive performance of PRS models built upon UKB-only and meta-analysis GWAS in the UK Biobank testing set, eMERGE and PAGE MEC cohorts. **A)** Prediction accuracy was measured using area under the curve (AUC) as a performance metric. Error bars indicate 95% confidence intervals of 10,000 non-parametric bootstrap replicates. Results were compared against AUC obtained with PRSs from Thompson et al. (2024) (pink line and shaded area as 95% CI). Conditions marked with an asterisk in green indicate those where our ensemble method performed significantly better at risk prediction. The condition marked with an asterisk in pink indicates a significantly better result for Thompson et al. DeLong’s test was used to compare the significant difference between AUC results. Performance for breast and prostate cancer was calculated using only female and male individuals, respectively. **B)** Similar to A), our PRS models were compared to Lennon et al. (2024) models in the eMERGE cohort. **C)** Prediction results summarized per PRS study and cohort. Relative percentage change in performance (AUC) in non-European groups compared to accuracy obtained in Europeans. **D**) Relative percentage change in performance (AUC) in eMERGE and PAGE MEC compared to accuracy obtained in the UK Biobank per ancestry group.

To validate the performance of our newly released PRS models, we applied them to the diverse cohorts of eMERGE and PAGE MEC, comprising 84,215 and 9,098 participants respectively. We extracted phenotypic data related to 30 clinical conditions from eMERGE and seven from PAGE MEC. Furthermore, we incorporated eight PRS models from Lennon et al. (2024) [16], that corresponded to the conditions investigated in this study and were specifically tailored for clinical implementation in diverse populations. We avoided using Lennon et al. models on the UKB testing cohort to prevent overfitting, as these models were trained with data that included the UKB dataset. Instead, we applied these models to the eMERGE cohort, replicating the same testing approach used in their original study. Our ensemble models significantly outperformed those by Lennon et al. for six out of eight traits (DeLong’s test for AUC, *P* < 0.05), achieving a median improvement in AUC of 1.97% (Fig. 3B).The UKB WBU summary statistics exhibited the poorest outcomes compared to other approaches for both eMERGE for the 30 traits and PAGE MEC for the seven traits, followed by better performances achieved through meta-analysis with SBayesRC and our ensemble models (Figs. 3C and S2).

To further evaluate PRS performance, we employed two additional metrics: odds ratios (OR) per standard deviation (SD) of PRS and a comparison between individuals in the top 20% of the PRS distribution, identified as high-risk, versus the remaining population. Consistent results with our primary findings were observed, as evidenced by a strong correlation between the AUC and these measures (Figs. S3 and S4). Specifically, our approach outperformed Thompson et al. [8] for 26 out of 30 traits using OR per SD as a metric and for 25 out of 30 traits using OR at the top 20%, and surpassed Lennon et al. [16] for seven out of eight traits using OR per SD as a performance metric and for eight out of eight using OR at the top 20%.

Subsequently, we analyzed the performance across different ancestries in UKB individuals, who were stratified by their genetically predominant ancestry using principal component analysis. In these analyses, we concentrate on populations that exhibit more than 50 cases of the trait being studied to ensure precise estimations. For eMERGE and PAGE MEC cohorts, we utilized the reported ethnicity labels for classification. Our findings were comparable to those of Thompson et al. [8] and Lennon et al. [16] (Fig. S5). As expected, we observed a decline in PRS accuracy for non-European ancestries. For instance, Latinos in eMERGE had a median decrease in AUC of 2.5%, while Africans experienced the largest reductions of 4.7% and 7.6% in eMERGE and UKB, respectively, compared to their European counterparts (Fig. 3D). An outlier were the East Asians in the UK Biobank, who exhibited only a minimal decrease in AUC of 1.3%, potentially reflecting the significant representation of Japanese samples from the BioBank Japan in our meta-analysis. Furthermore, we evaluated the impact of applying our ensemble PRS models, developed on UK Biobank data, to other cohorts. Comparing performance by ancestry for each trait within the UK Biobank to results in eMERGE or PAGE MEC, we found expected performance drops, but with none exceeding a 4% median reduction—specifically 3.6% and 3.2% for Asians and Europeans, respectively, while no drop for Africans was reported (Fig. 3E).

In evaluating the contributions of each PRS algorithm within our ensemble framework, SBayesRC emerged as the most influential component, accounting for 43.7% of the ensemble weight on average. This was followed by SCT and LDpred2, which were assigned weights of 17.3% and 15.7%, respectively. Algorithms such as PRS-CS and lassosum2 had comparatively lower weights of 12.5% and 10.8%, respectively. Notably, the allocation of weights to the algorithms closely aligned with their performance rankings observed in our preliminary benchmarking assessments.

### Addition of Ancestry and Clinical Characteristics to PRS Models

We proceeded to enhance our ensemble PRS models by incorporating additional information, initially integrating ancestry details. We applied an iterative approach, settling on the use of a combination of ancestry information as deduced by Orchestra [82], a local ancestry inference (LAI) software, in addition to the traditional first four genotyping principal components (PCs). We trained a model that combines PRSs with ancestry information using logistic regression for each trait, utilizing the same 30,000 participants previously employed for retraining our ensemble model. This approach led to a marginal improvement in overall accuracy (Fig. 4A and B). Specifically, adding only the PC components to the baseline (PRS-alone) model enhanced performance for 24 out of 30 traits analyzed, with a median AUC improvement of 0.3%. Moreover, the addition of detailed ancestry information from Orchestra resulted in accuracy gains for 22 traits, providing an additional median AUC enhancement of 0.1% beyond the addition of PCs alone. Collectively, ancestry information improved accuracy estimates for 26 traits, a significant albeit subtle enhancement (*P* = 0.00006, sign test).

**Fig. 4.**
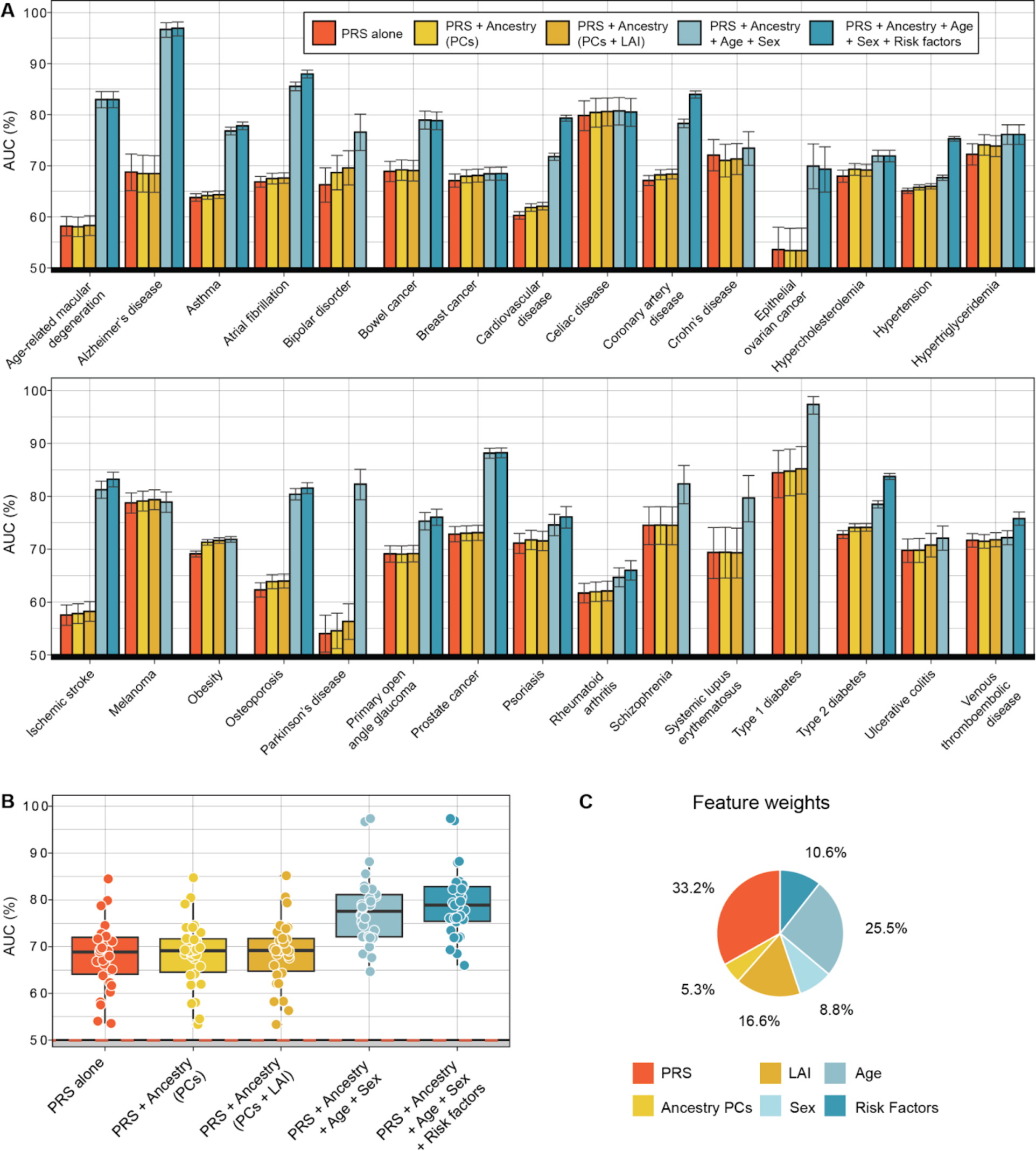
Predictive performance of risk models incorporating ancestry, age, sex and risk factors information. **A)** Prediction accuracy was measured using AUC in the testing set of UKB. Bars indicate 95% confidence intervals of 10,000 non-parametric bootstrap replicates. Models shown include performance with: ensemble PRS alone; genotyping PCs for ancestry information; Orchestra for local ancestry deconvolution; and additional factors such as sex, age, and relevant risk factors. **B)** Prediction results summarized per risk model where each dot represents a clinical condition. If a trait lacked relevant risk factors, the accuracy of the final comprehensive model equaled that of the preceding model, which included only prs, ancestry, age and sex **C)** Averaged weights per feature in the final logistic regression model across 30 studied traits.

While creating predictive risk models solely based on genetic information is a good way to test and benchmark various PRS models against each other, in a real world clinical setting, the addition of non-genetic information to the final model is needed in order to increase performance. With this in mind, we incorporated other easily obtained clinical characteristics. We added this information to PRS and ancestry to make a final logistic regression model that would serve as a prospective diagnostic test, with an easily interpretable positive or negative value, typical of other diagnostic tests used in a clinical setting. Table S3 contains the various inputs for each trait model which includes additional variables: sex, age and risk factors. In this regard, there was improvement in a large subset of the traits tested with 12 models surpassing the 80% accuracy range with the AUC metric. Including age and sex to the previous model improved performance for all but one trait, with a median AUC improvement of 8.4%. The addition of risk factors (if there were any) added a modest extra 1.3% on average to the final performance. Specifically, 16 out of 21 conditions with defined risk factors improved their accuracy mark (*P* = 0.03, sign test). Overall, incorporating additional factors relevant to disease risk prediction consistently yielded notable enhancements.

When we look at the contributions of individual features to the full risk models, PRS proved to be the predominant factor, contributing roughly one-third of the predictive weight on average (Fig. 4C). Although the addition of ancestry information to the models did not substantially enhance performance over PRS alone, up to 22% of the model’s weight was attributable to ancestry data, suggesting that our PRS model may have partially captured some ancestry effects. Age was second only to genetics, accounting for an average of 25% of the model’s weight. Gender and other risk factors were less influential, contributing 8.8% and 10.6% respectively.

Significant variability was evident across traits (Fig. S6). For instance, conditions such as Alzheimer’s disease (74% of total weight), age-related macular degeneration (71%), Parkinson’s disease (64%), and prostate cancer (51%), which predominantly manifest later in life, showed a higher weighting for age. Conditions where there is a known gender bias had a higher weighting for gender, e.g. systemic lupus erythematosus (37% of total weight), osteoporosis (32%), total cholesterol (29%), rheumatoid arthritis (28%), and coronary artery disease (20%). Notably women are more commonly affected by lupus, osteoporosis, and rheumatoid arthritis, while men tend to have higher cholesterol levels and more frequently suffer from coronary artery disease. Cardiovascular-related conditions such as hypertension and coronary artery disease, along with other conditions like venous thromboembolism, atrial fibrillation, and ischemic stroke, displayed a high dependency on risk factors such as body mass index, pre-existing hypertension and prior cardiovascular events. Type 2 diabetes was impacted by body mass index.

Several clinical conditions demonstrated a substantial reliance on ancestry. Examples include Crohn’s disease (93% total weight), celiac disease (60%), and type 1 diabetes (47%), which are know to be more prevalent in European populations; ischemic stroke (37%) and schizophrenia (31%), which exhibit higher rates in African-American populations; or type 2 diabetes (30%), which has a higher prevalence in non-Europeans. Melanoma, bipolar disorder, and breast cancer were the diseases most affected by genetics (PRS) in our models. Particularly, the melanoma risk model was almost entirely based on PRS, achieving an accuracy of nearly 80% with PRS alone.

### Towards the Clinical Implementation of PRS-Based Disease Risk Models

Measuring PRSs early on in one’s life offers a window into future health risks and identifies individuals who might face medical challenges as they age. Evidence from cumulative incidence curves, including the UK Biobank (Fig. S7) and additional cohorts like eMERGE and PAGE MEC (Fig. S8), supports the notion that higher PRSs correlates with a lifelong increased probability of developing health conditions. However, current health systems prioritize rare mutations that confer significant risks for disease development, while neglecting PRSs that can aggregate comparable risks through the accumulation of hundreds to thousands of common variants with minor effects. Indeed, for numerous prevalent diseases, certain genes have been discovered where rare mutations significantly increase the risk, often several-fold, for carriers who are heterozygous [85]. In line with Thompson et al. (2024) [8], we analyzed and contrasted the risk profiles of rare mutations and PRSs, using cardiovascular disease, breast cancer and bowel cancer as illustrative case studies.

In our analysis of UKB participants, we identified carriers of high-risk mutations in key functional genes via whole exome sequencing and aimed to match their risk profiles by selecting individuals within the top percentiles of higher risk based on their PRS profiles. For example, the prevalence of coronary artery disease among carriers of pathogenic mutations in genes associated with familial hypercholesterolemia (*APOB, APOE, LDLR and PCSK9*) was approximately 12% by age 70 in our testing cohort. A similar lifetime risk was noted among participants in the top 20% of the PRS distribution outlined by Thompson et al. (2024) [8] for this trait, consistent with what they reported. Our PRS ensemble model identified even more individuals – up to 22% of those in the top distribution – as having an equivalently high risk (Fig. 5A). Remarkably, the high PRS group identified 55 to 80 times more true coronary artery disease events than those detected among carriers of high-risk variants, depending on the age group analyzed (Fig. 5B). This incidence surpassed the one reported by Thompson et al., which identified between 50 and 73 times the cases found in carriers. Rare pathogenic mutations contribute more significantly to early-onset disease, evidenced by a lower ratio of high PRS individuals to rare mutation carriers among those diagnosed before age 50. We also explored the interplay between monogenic risk variants and PRS profiles, categorizing individuals into low, intermediate, or high PRS risk groups. The risk among mutation carriers, compared to non-carriers with an intermediate PRS, varied from a 1.22-fold odds ratio in the lowest PRS risk group to 7.05 in the highest PRS risk group (Fig. 5C). This underscores the critical need to consider all genetic components and illustrates how PRS can modulate the effects of high-risk variants typically evaluated in clinical settings. Parallel insights emerge when examining breast cancer in relation to mutations in *BRCA1*, *BRCA2, ATM, PALB2*, and *CHECK2*, and bowel cancer associated with rare variants in *MSH2, MSH6, MLH1*, and *PMS2* (Fig. S9).

**Fig. 5.**
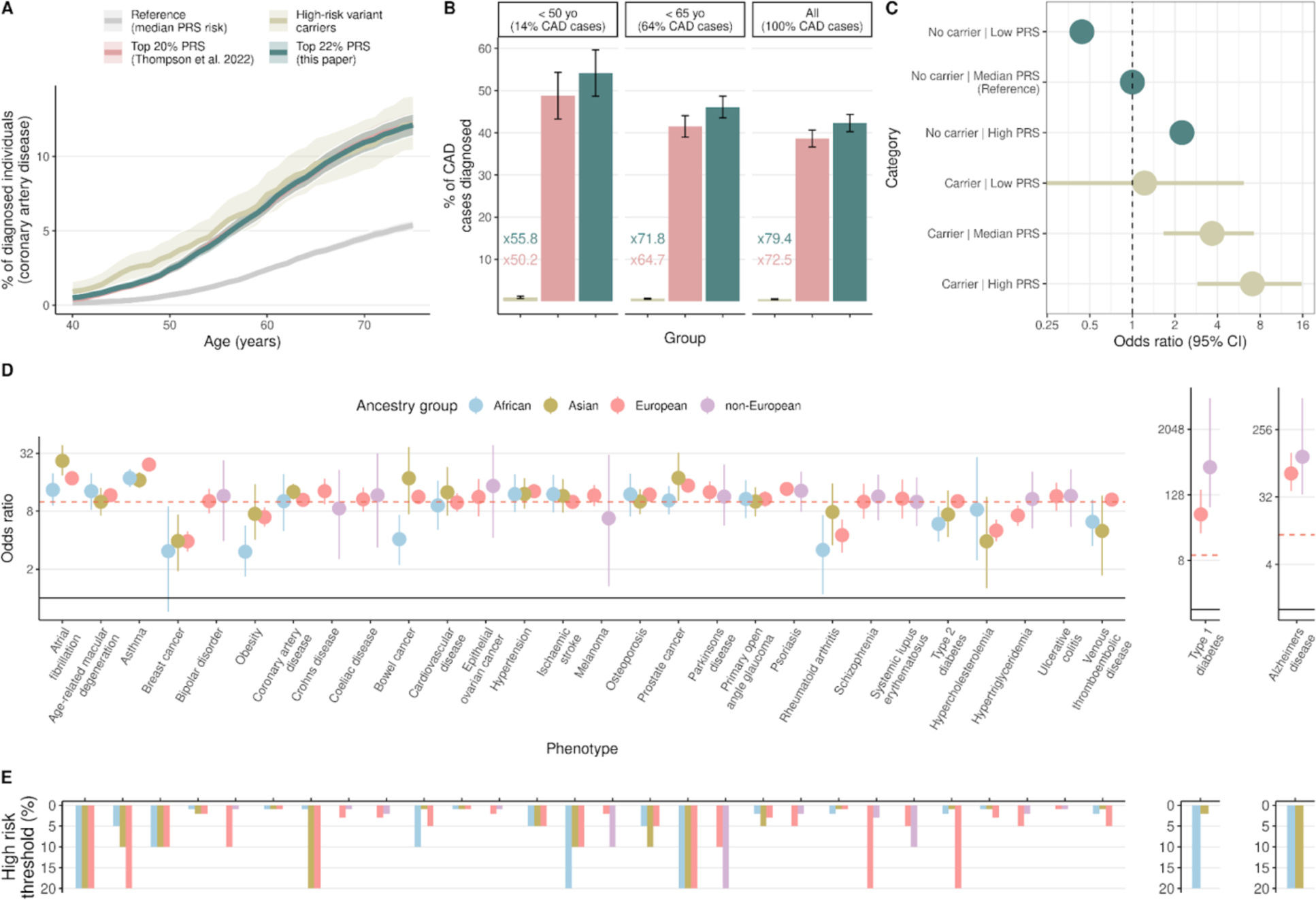
Towards the clinical implementation of PRS-based models. Comparison between our PRS risk profiles and functional variant carriers in UKB Europeans. **A)** Cumulative incidence of coronary artery disease among carriers with pathogenic or likely-pathogenic mutations in familial hypercholesterolemia genes vs. individuals in the top 22% of our PRS distribution from the ensemble method, where a percentile was chosen to match the risk up to age 70 with that of mutation carriers. Additionally, the median 25%-75% of the PRS distribution serves as the population reference. **B)** Percentage of coronary artery disease (CAD) cases diagnosed in individuals aged under 50, under 65, and across all age groups, occurring either in mutation carriers or in those at the top of the PRS distribution. The plots display the ratio of high PRS cases to mutation carrier cases in each age category. **C)** The forest plot displayed odds ratios for the observed risk in coronary artery disease, with horizontal lines representing the 95% confidence intervals. Individuals were categorized into three groups based on their polygenic scores – low, intermediate, and high, corresponding to the lowest 25%, middle 25-75%, and highest 25% of the distribution, respectively. The intermediate polygenic score group served as the reference category for calculating odds ratios. **Diagnostic test effectiveness across medical traits. D)** Odds ratios are reported per ancestry group, defined as the ratio of the odds that an individual with a clinical condition will score above a specified risk threshold, to the odds of scoring below this threshold without the condition. Error bars represent 95% confidence intervals. **E)** ‘High risk threshold’ illustrates the percentile cutoff for a specific condition, above which results are considered high-risk. Odds ratios were optimized using thresholds at the top 2%, 3%, 5%, 10%, and 20% percentiles.

Many of the risk models we have developed in the previous section demonstrated sufficient overall accuracy to potentially merit being used in a clinical setting. To further elucidate the effectiveness of these models as diagnostic tests, we next calculated the diagnostic odds ratio (DOR) [86] for each model. To achieve sufficient sample sizes for certain traits, we aggregated East Asians and South Asians from the UK Biobank into a broader ‘Asian’ category. If non-European populations did not meet this threshold, we combine all non-European groups (African and Asian) into a ‘Non-European’ category for the analysis. We adjust the odds ratios to exceed a diagnostic odds ratio of 10, while simultaneously maximizing the number of individuals in the population who are at least at this risk level, by focusing on the top 1%, 2%, 3%, 5%, 10%, or 20% of the population at higher risk. As can be seen in Fig. 5D and E, diagnostic odds ratios ranged from a low of 3.04 for obesity in Africans to a high of 411.67 for type 1 diabetes in non-Europeans. However, when we restrict our observations to condition-ancestry pairs that have over 100 cases, the highest DOR of 66.2 was observed for Alzheimer’s disease in Europeans. The DOR across all ancestry groups exceeded 5 for 25 out of 30 traits and 10 for 19 out of 30 traits, suggesting that the majority of the tests were highly predictive.

## Discussion

Polygenic risk scores (PRS) represent a new frontier in the field of personalized medicine, with the potential to predict an individual’s susceptibility to various diseases by aggregating the effects of numerous genetic variants across the genome. The promise of PRS lies in their ability to harness the wealth of data generated by genome-wide association studies (GWAS), thereby enabling risk stratification at a population level and the possibility of targeted interventions for those at high risk [1-3,6,7,9]. This could lead to more precise preventive measures, refined screening protocols, and tailored therapeutic strategies, ultimately improving clinical outcomes. However, the utility of PRS is currently hampered by several limitations. Chief among these is the reduced predictive power in diverse populations due to the majority of GWAS being conducted in individuals of European ancestry, leading to biased risk predictions when applied to other ethnic groups [1,8]. Additionally, the integration of PRS into clinical practice faces challenges, including the need for validated models that have the same level of accuracy and utility of traditional diagnostic tests used in a clinical setting [9,10].

We generated novel genome-wide summary statistics for 30 medically-related traits, leveraging trans-ancestry genome-wide association study (GWAS) meta-analyses of diverse cohort data, including datasets such as the UKB, the FinnGen project, BioBank Japan, and the Global Lipids Genetics Consortium. Using meta-analyses often yielded notable improvements in PRS accuracy over UKB only. We believe there are two main reasons for that: (1) the meta-analyses had a higher number of cases compared to UKB, on average 6.4 times greater for binary traits and 7 times greater for quantitative traits; and in addition (2) the meta-analyses increased the representation of non-European ancestries where such data was available.Trait-association resolution is often enhanced by the inclusion of diverse ancestries, an effect attributed to heterogeneity in allele frequencies and linkage disequilibrium, which may have aided the mapping of causal variants and improved PRS accuracy across populations [87]. However, challenges like data harmonization remain, emphasizing the need for better integration practices as more datasets become available.

Among the six tested PRS models, SBayesRC had the overall best performance. Leveraging functional genomic annotations in SBayesRC may play a critical role in distinguishing causal from non-causal variants, further enhancing performance alongside increasing GWAS sample size and diversity [77]. One key advantage of this model may be that it is the only one to exclusively utilize summary statistics from GWAS without depending on a training cohort with individual-level data for tuning hyper-parameters. In contrast, many PRS methods still require tuning parameters via grid searches on external datasets, which can compromise statistical power by requiring split testing samples. It is important to note, however, that SBayesRC and PRS-CS use UKB as an LD reference panel. It is possible that this may inadvertently inflate the results in UKB, and may be partially responsible for the drop in accuracy when applied to eMERGE and PAGE MEC datasets. However, a meta-analysis incorporating multiethnic datasets would likely alter the LD configuration compared to UKB alone. Furthermore, despite PRS-CS also using UKB LD as a reference, it still did not achieve the same accuracy levels as SBayesRC.

To maximize the accuracy of our PRSs, we trained an ensemble PRS model using logistic regression combining the outcomes from five PRS algorithms. Our ensemble model performed better than the SBayesRC algorithm for 23 out of 30 traits in our UKB testing cohort. Moreover, our results proved superior in AUC and OR per SD to those reported by Thompson et al. [8] for 26 out of 30 traits, demonstrating significant improvements in AUC for 22 of these traits, where our median AUC was 5.07% higher. On eMERGE, our ensemble models significantly outperformed those by Lennon et al. [16] for six out of eight traits when using AUC as a metric, for seven out of eight traits when it comes to OR per SD, and for all eight traits when we measured OR at the top 20%. These improvements can likely be partially attributed to larger sample sizes incorporated into our meta-analysis and more diverse datasets included in the meta-analyses, with further gains achieved by using our ensemble PRS model. When it comes to sample size, specifically, our study achieved a 1.25-fold increase in controls and a 1.22-fold increase in cases compared to Thompson et al. Twenty-one out of 27 binary diseases for controls and 22 out of the 30 clinical conditions for cases showed superior numbers, notwithstanding some overlap in the cohorts meta-analyzed. Only three traits, epithelial ovarian cancer, Alzheimer’s disease and breast cancer, had fewer cases and controls simultaneously compared to those reported by Thompson et al. In the case of epithelial ovarian cancer, the results were particularly poor within our UKB validation cohort. For Alzheimer’s disease and breast cancer, although our initial outcomes were weaker, the application of our ensemble method yielded improvements that rivaled Thompson et al.’s results. Indeed, our ensemble approach proved efficient by training PRS algorithms on a computationally manageable cohort and then combining them within a larger training dataset, optimizing the computational workflow while achieving greater accuracy, an approach made feasible by the simplicity and scalability of logistic regression. Altogether, by surpassing the models of Thompson et al. and Lennon et al., which had already been optimized and outperformed numerous published PRS models, our findings demonstrate that our PRS models are moving remarkably toward clinical application.

It is important to note that methodological differences between our study and those cited, such as variations in phenotype definitions and the cohorts used for evaluation, may contribute to discrepancies in published accuracy metrics. For instance, the results by Lennon et al. [16] were derived from a smaller eMERGE cohort consisting of 2,500 individuals, whereas our evaluation utilized a substantially larger sample from the same project. In this regard, AUC metric tends to be more robust with larger sample sizes, an essential factor particularly in contexts of conditions with low prevalence. Although this likely contributed to the minor discrepancies in accuracy metrics reported across the studies, the significance of our findings remains robust.

When applying our ensemble PRS models, developed on UKB data, to eMERGE or PAGE MEC cohorts, the drop in performance did not exceed a 4% median reduction. This suggests that our ensemble PRS model was well calibrated. As previously noted [88], we observed a decline in PRS accuracy for non-European ancestries. Africans experienced the largest reductions in accuracy compared to their European counterparts, while East Asians showed the smallest decrease, likely due to the representation of BioBank Japan samples in our meta-analysis. This again underscores the importance of including diverse ethnic groups in PRS training to enhance its global applicability.

Next we incorporated ancestry information and other easily obtained clinical characteristics: sex, age and known risk factors, to make a final logistic regression model that would serve as a prospective diagnostic test. This resulted in an improvement in a large subset of the traits tested, with 12 models surpassing the 80% accuracy range with the AUC metric. Age and sex improved performance for all but one trait, with a median AUC improvement of 8.4%. Addition of risk factors (where available) added an extra 1.3% on average to the final performance, while ancestry information improved accuracy by a modest 0.4% on average. Although the addition of ancestry information to the models did not substantially enhance performance over PRS alone, when we look at the contributions of individual features to the full risk model, up to 22% of the model’s weight was attributable to ancestry data. This suggests that our PRS ensemble model might have implicitly captured the effects of ancestry, potentially through the integration of various PRS models tailored to different ancestries. Notably, local ancestry inference had a more pronounced contribution than traditional PCs, suggesting that detailed ancestry insights might lead to greater predictive accuracy.

It is important to highlight the variability in the contributions of individual features to the model that was evident across traits. We observed higher contribution of genetics (PRS) to models such as melanoma, age to models such as Azhaimer’s disease, gender to models such as systemic lupus erythematosus, ancestry to models such as Crohn’s disease and known risk factors to models such as coronary artery disease. We can conclude that depending on the condition, incorporating easily measured clinical factors alongside genetic data into predictive models, can be an easy way to increase model accuracies, even without considering variables such as smoking status or lifestyle.

Finally, we compared the predictive accuracy of our PRS models to that obtained by looking at rare pathogenic variants, for three well studied conditions, coronary artery disease, breast cancer and bowel cancer. For coronary artery disease, our PRS model was able to identify between 55 and 80 times more true coronary artery disease events than models using rare pathogenic variants. Rare pathogenic mutations seemed to contribute more to early-onset disease, while more common genetic variants with modest effects (found in our PRS) seemed to capture relatively more late-onset cases, in line with Thopson et al. [8]. When we look at the interplay between rare pathogenic risk variants and PRS profiles, the risk among rare pathogenic mutation carriers varied from a 1.22-fold odds ratio in the lowest PRS risk group to 7.05 in the highest PRS risk group. Similar patterns were observed for breast and bowel cancer. This underscores the critical need to consider all genetic components and further suggests that the polygenic component can modulate the effects of high-risk variants typically evaluated in clinical settings. The diagnostic odds ratios across all ancestry groups exceed 5 for 25 out of 30 traits and 10 for 19 out of 30 traits, suggesting that the majority of the tests were highly predictive.

In this paper, we have optimized PRS models both by increasing GWAS power through meta-analysis and by using ensemble models that leverage the best features of individual PRS models. Further, we showed the importance of integrating a variety of data types to systematically improve diagnostic accuracy. We demonstrate that many of our models have sufficient accuracy to warrant consideration of being used in a clinical setting. A next step would be to fully validate the entire process from instrumental genotype measurements to final classification in individuals that do not belong to cohorts used for training and validation in this paper. With ever increasing GWAS sample sizes, refinements, and improvements in PRS algorithms, we expect that the models will further improve with time. As our current benchmarking demonstrates, we are one step closer to using PRS in a clinical setting across various ancestral populations.

## Data and code availability

This study used the openly available GWAS catalog, FinnGen and deCODE genetics datasets and the UK Biobank, dbGaP and The Million Veterans Program datasets that are available to researchers upon application. UK Biobank research was conducted under application number #84038. dbGaP access was obtained for phs001868.v1.p1 (Landi et al., 2020), phs001584.v2.p2 (eMERGE cohort), phs000220.v2.p2 (PAGE MEC cohort) and phs001672.v11.p1 (The Million Veterans Program). All data produced in the present study are available upon request to the corresponding author.

## Acknowledgments

We thank the team and our colleagues at Omics Edge and Genius Labs.

## Author contributions

J.L.J. and P.G.Y. designed this study. J.L.J, S.B. and A.D.M. processed genomic data for training and testing. J.L.J, A.T. and B.N. gathered and processed summary statistics for the meta-analyses. J.L.J., A.T. and M.K. performed the meta-analyses. J.L.J., A.T., A.O., and C.M. developed the integrated PRS disease prediction models. J.L.J., B.N. and P.G.Y. wrote the main manuscript text. J.L.J. prepared the figures. All authors read and approved the final manuscript.

## Declaration of Interests

J.L.J., A.T., B.N., A.O., C.M., S.B., A.D.M. and P.G.Y. are either employed by and/or hold stock or stock options in Omics Edge, a subsidiary of Genius Labs. In addition, P.G.Y. has equity in Systomic Health LLC and Ethobiotics LLC. This does not alter our adherence to journal policies on sharing data and materials. There are no other relevant activities or financial relationships which have influenced this work.

All work was funded by a commercial source, Omics Edge, a subsidiary of Genius Labs Company. Omics Edge provided only funding for the study, but had no additional role in study design, data collection and analysis, decision to publish or preparation of the manuscript beyond the funding of the contributors salaries.

## Web resources

R Project, https://www.R-project.org/

GWAS catalog: https://www.ebi.ac.uk/gwas/

FinnGen: https://www.finngen.fi/en

deCODE genetics: https://www.decode.com/

UK Biobank: https://www.ukbiobank.ac.uk/

dbGaP (PAGE MEC): https://www.ncbi.nlm.nih.gov/projects/gap/cgi-bin/study.cgi?study_id=phs000220.v2.p2

dbGAP (eMERGE): https://www.ncbi.nlm.nih.gov/projects/gap/cgi-bin/study.cgi?study_id=phs001584.v2.p2

dbGAP (The Million Veterans Program): https://www.ncbi.nlm.nih.gov/projects/gap/cgi-bin/study.cgi?study_id=phs001672.v11.p1

## Notes

### Competing Interest Statement

J.L.J, A.O., C.M., A.T, B.N., S.B., A.D.M. and P.G.Y. are either employed by and/or hold stock or stock options in Omics Edge, a subsidiary of Genius Labs. In addition, P.G.Y. has equity in Systomic Health LLC and Ethobiotics LLC. This does not alter our adherence to journal policies on sharing data and materials. There are no other relevant activities or financial relationships which have influenced this work.

### Author Declarations

This study used the openly available GWAS catalog (https://www.ebi.ac.uk/gwas/home), Finngen (https://www.finngen.fi/en) and deCODE genetics (https://www.decode.com/summarydata/) datasets and the UK Biobank (https://www.ukbiobank.ac.uk/), dbGaP (https://www.ncbi.nlm.nih.gov/projects/gap/cgi-bin/study.cgi?study_id=phs001868.v1.p1) and The Million Veterans Program (https://www.ncbi.nlm.nih.gov/projects/gap/cgi-bin/study.cgi?study_id=phs001672.v11.p1) datasets that are available to researchers upon application. UK Biobank research was conducted under application number #84038 and The Million Veterans Program #30221. dbGaP access was obtained for phs001868.v1.p1.

### Summary of Updates

- 2 more PRS algorithms were benchmarked, in addition to the 4 in version 1; - an ensamble model was developed using logistic regression to combine outputs from 5 top-performing algorithms; - the ensamble model was validated in two additional datasets: PAGE MEC and eMERGE; - the performance of 3 of the final disease prediction models was compared to the predictive performance of high-impact rare pathogenic variants; - Text, figures and supplemental files have been updated to reflect these changes.

