## Supplementary Figures and Tables for "Optimization of Multi-Ancestry Polygenic Risk Score Disease Prediction Models"

### Supplemental Figures and Tables

#### Supplemental figures and legends

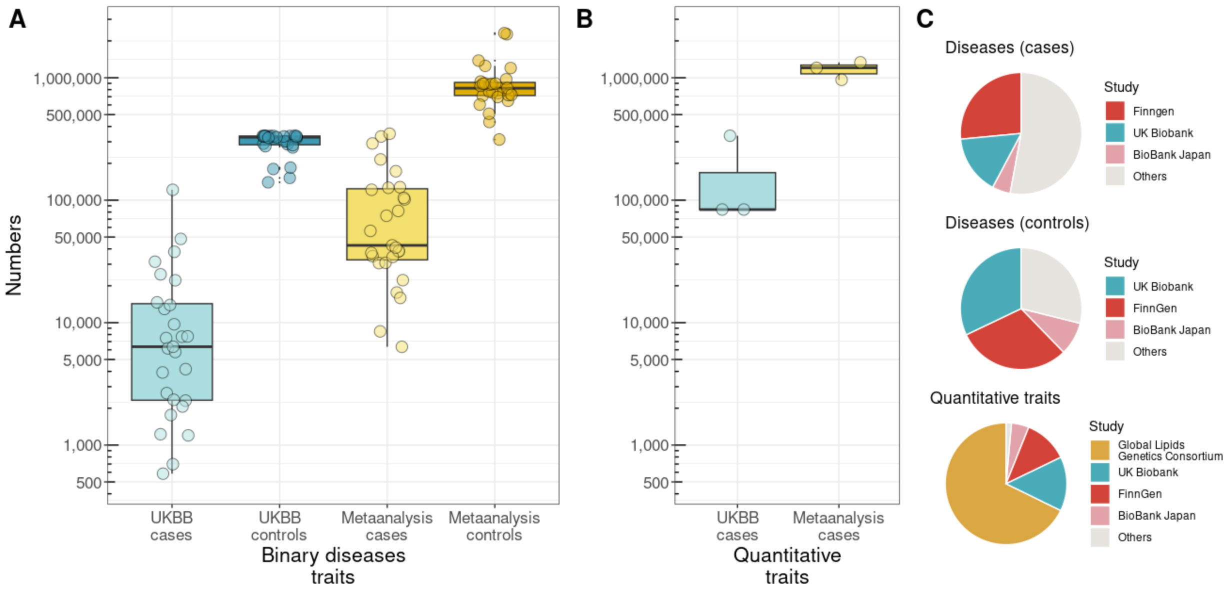


**Fig S1. Sample size and main studies in the meta-analysis. A)** Number of cases and controls for binary traits, comparing the UKBB summary statistics and the meta-analysis. Each dot represents a single trait. **B)** Sample sizes for quantitative traits, with all individuals treated as cases due to the PRS models being trained on quantitative measures. **C)** Proportion of samples per source, highlighting the four largest contributors to the meta-analysis.

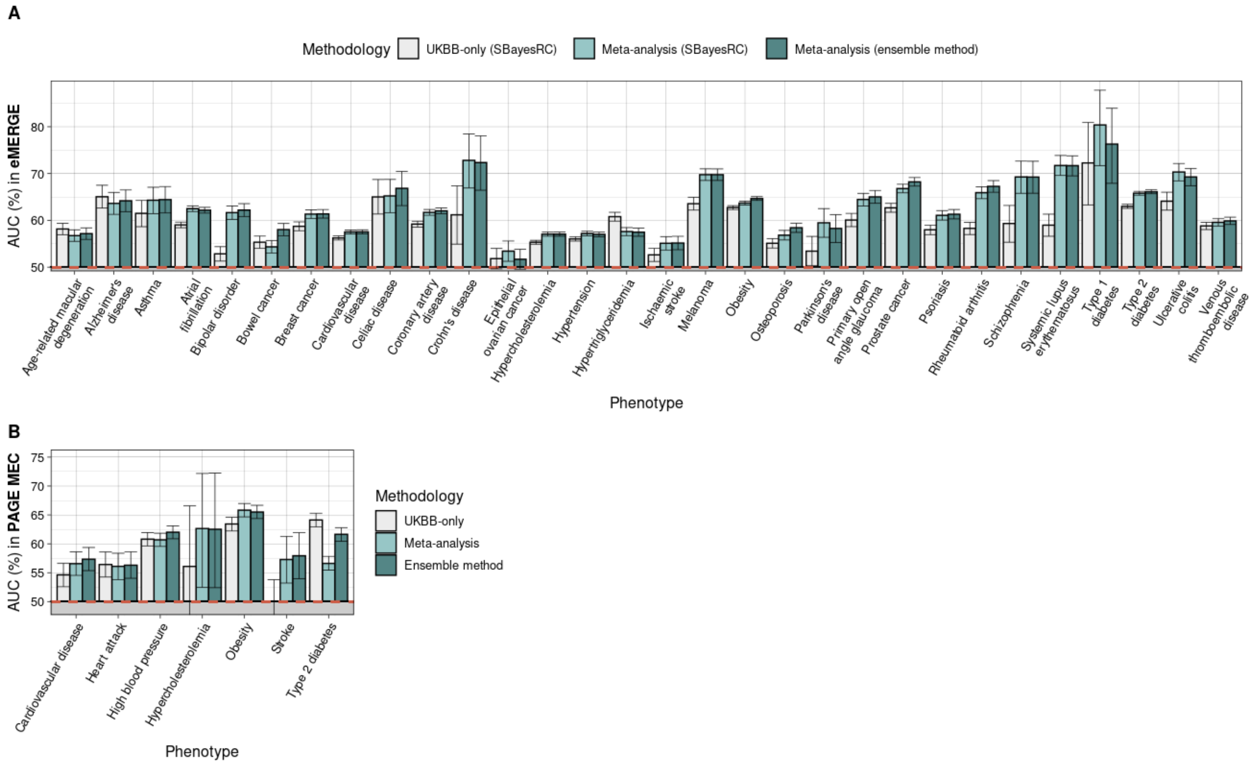


**Fig S2. Predictive performance of PRS models built upon UKB-only and meta-analysis GWAS.** Prediction accuracy was measured using area under the curve (AUC) as a performance metric for eMERGE **(A)** and PAGE MEC **(B)** cohorts. Error bars indicate 95% confidence intervals of 10,000 non-parametric bootstrap replicates. Performance for breast and prostate cancer was calculated using only female and male individuals, respectively.

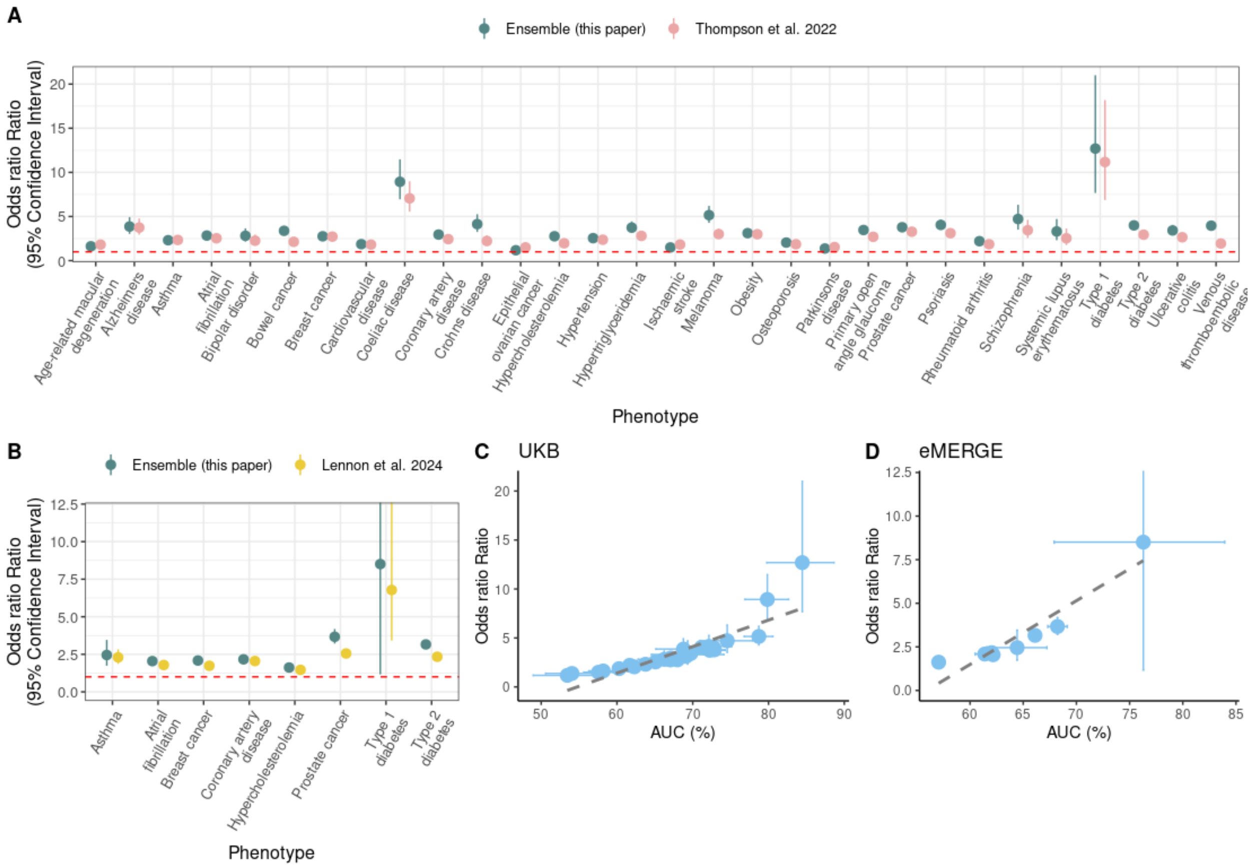


**Fig S3. Predictive performance of PRS models comparing our profiles from our ensemble method and results by other studies using odds ratios as a performance metric. A)** Prediction accuracy was measured using odds ratio by comparing individuals in the top 20% of the PRS distribution, identified as high-risk, versus the remaining population. Results were compared against those obtained with PRSs from Thompson et al. (2024). Error bars indicate 95% confidence intervals (binomial test). **B)** Same as A) but our PRS models and Lennon et al. (2024) models were employed on the eMERGE cohort. **C), D)** Correlation between odds ratios and AUC (%) across all traits tested both in UKB **(C)** and eMERGE **(D)** cohorts.

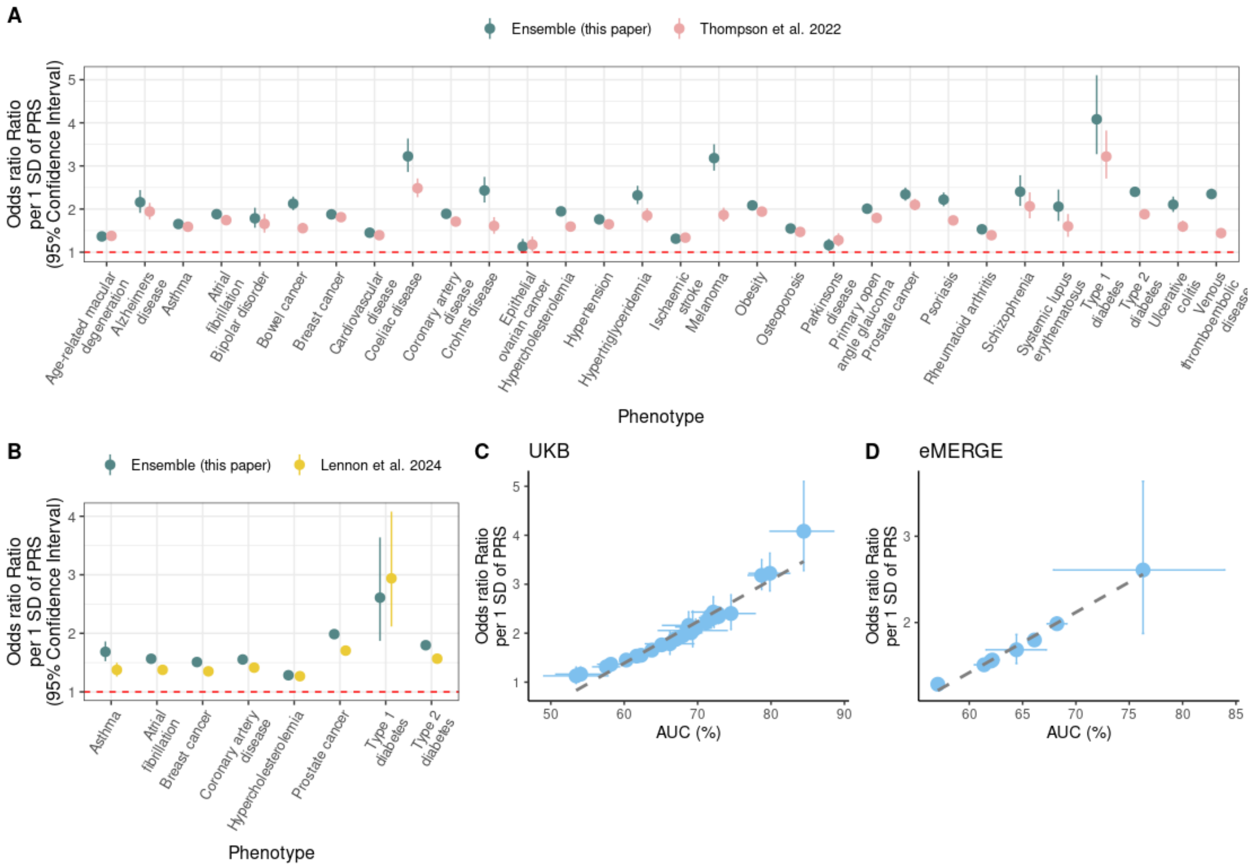


**Fig S4. Predictive performance of PRS models comparing our profiles from our ensemble method and results by other studies using odds ratios per standard deviation of PRS as a performance metric. A)** Prediction accuracy was measured as done in Thompson et al. (2024) article. Results were compared against those obtained with PRSs from Thompson et al. (2024). Error bars indicate 95% confidence intervals (binomial test). **B)** Same as A) but our PRS models and Lennon et al. (2024) models were employed on the eMERGE cohort. **C), D)** Correlation between odds ratios and AUC (%) across all traits tested both in UKB **(C)** and eMERGE **(D)** cohorts.

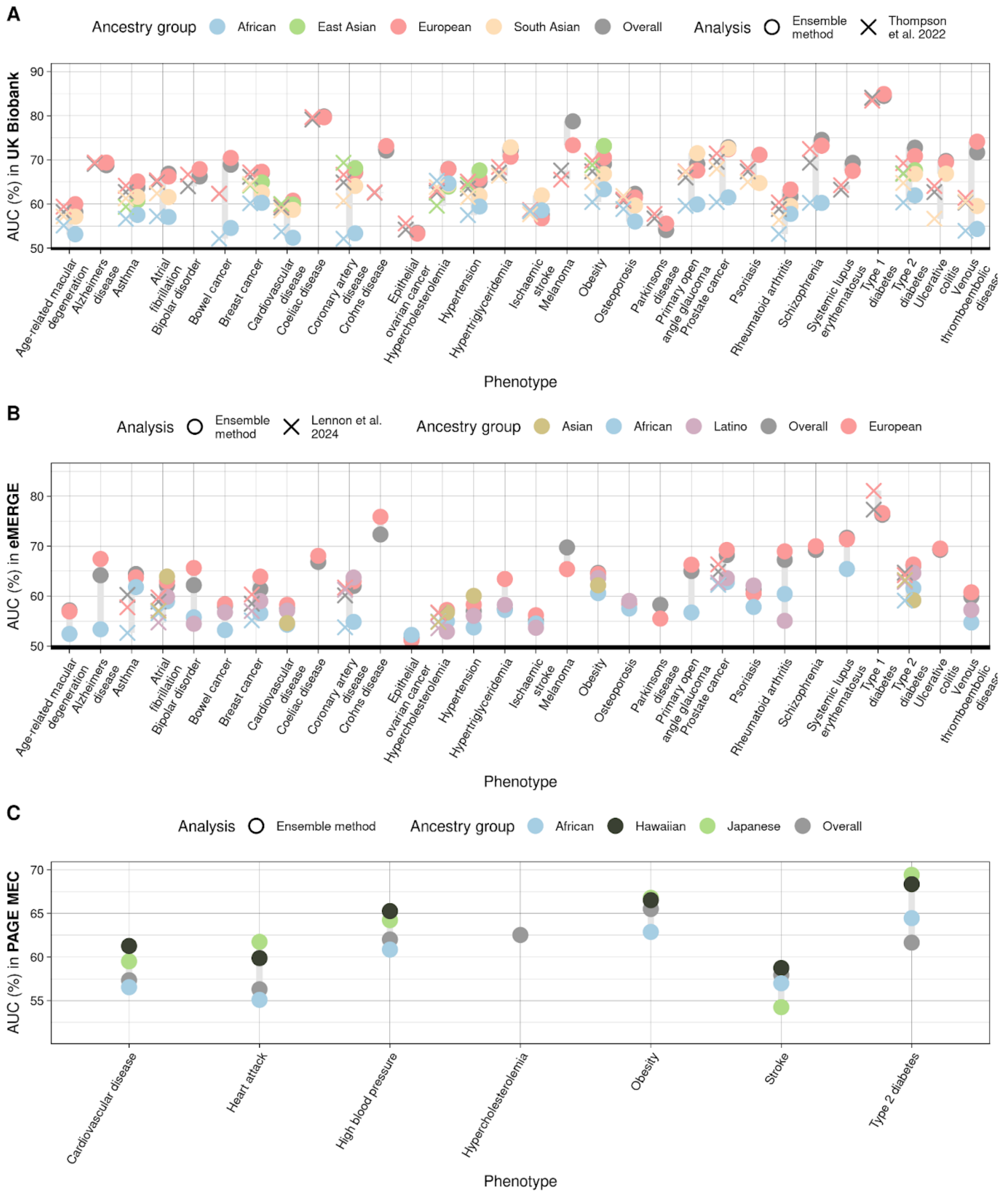


**Fig S5. Predictive performance of our PRS models per ancestry built upon meta-analysis GWAS in the UK Biobank testing set with our PRS ensemble method.** Prediction accuracy was measured using area under the curve (AUC) as a performance metric. Our findings were comparable to those of Thompson et al. (2024) and Lennon et al. (2024). Only ancestry groups with at least 50 cases for the clinical condition were included, with the exception of Europeans with type 1 diabetes, which is a low prevalence disease but highly predictable according to the AUC metric.

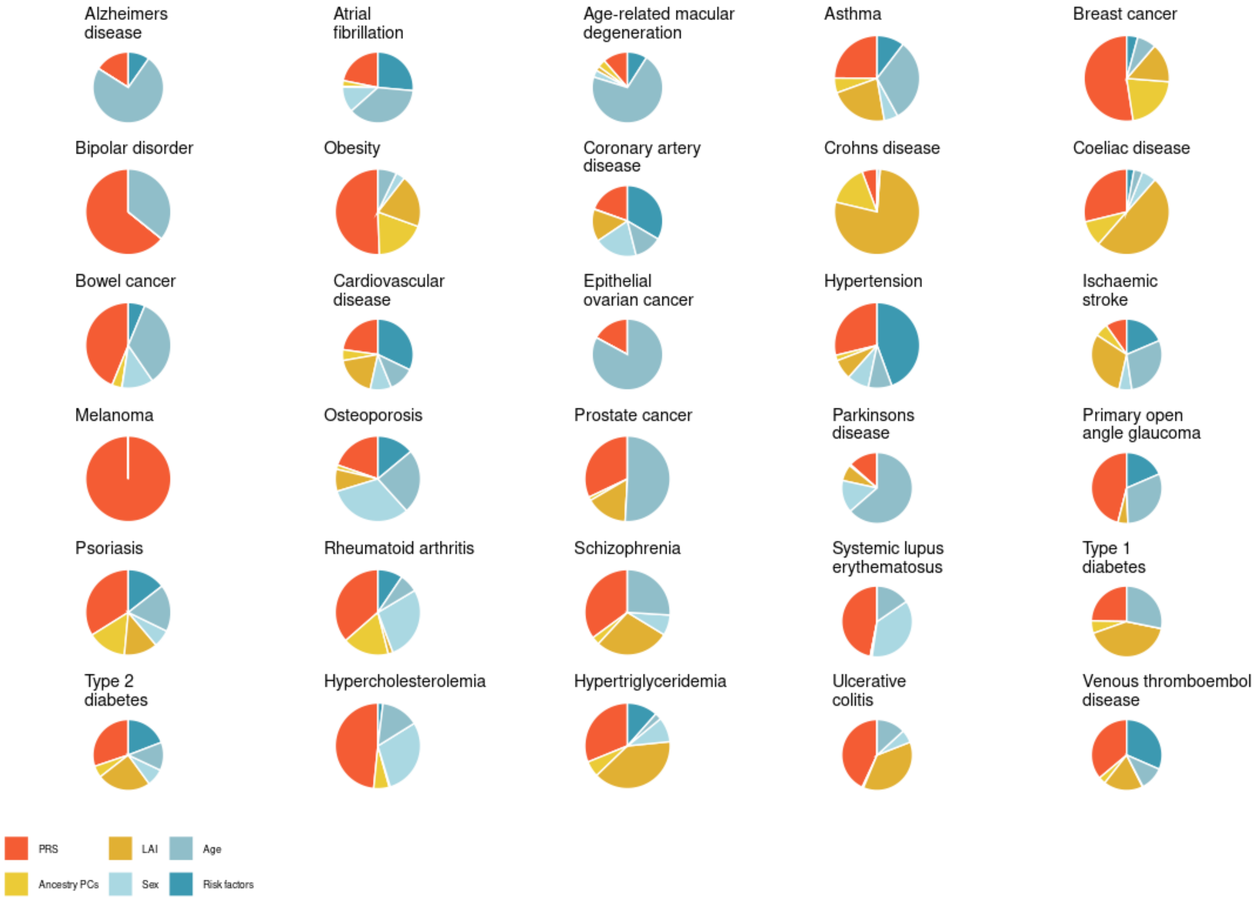


**Fig S6. Weights per feature in the final logistic regression model for all the 30 studied traits.**

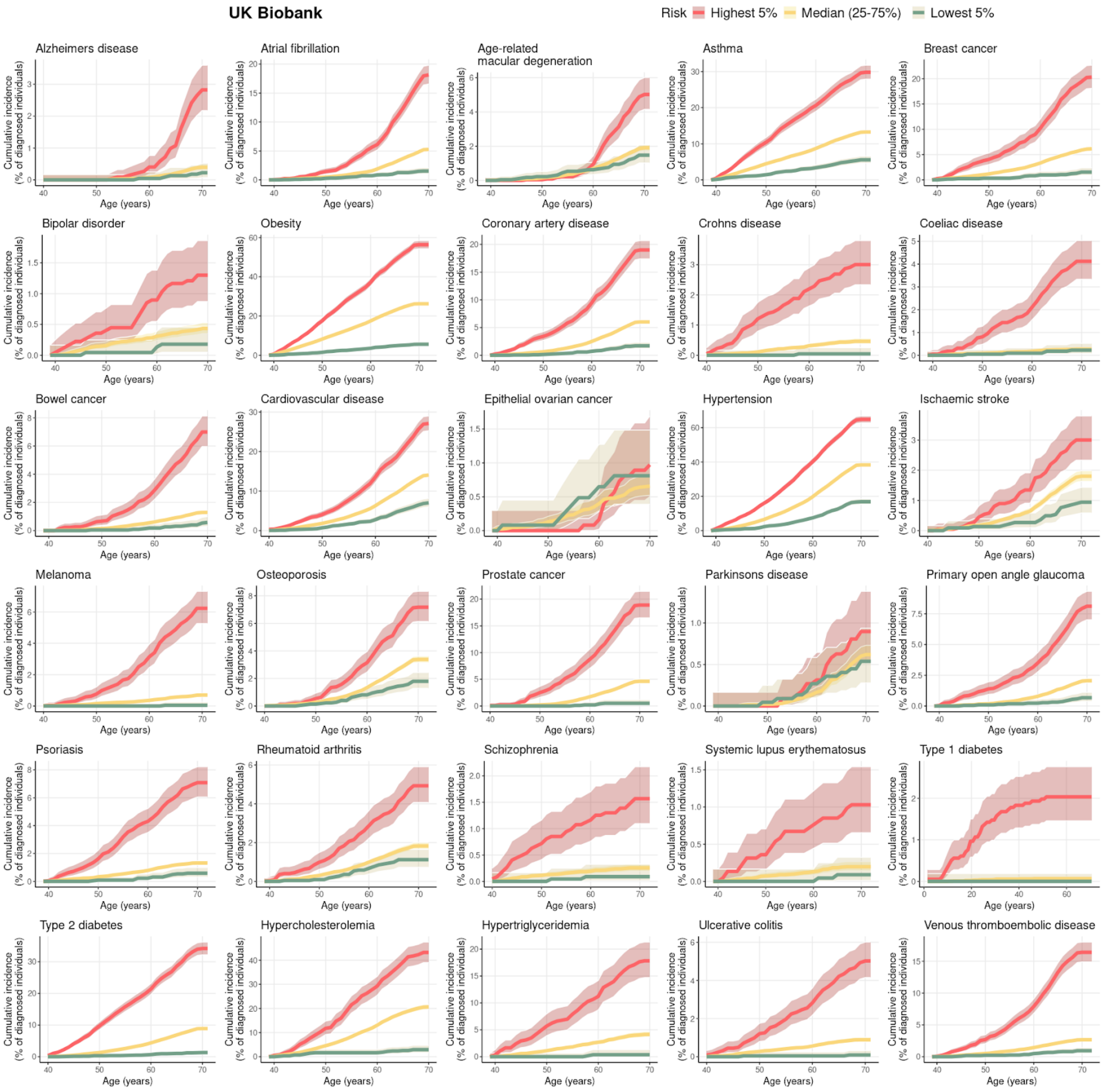


**Fig S7. Cumulative incidence graphs display the predictive accuracy of our polygenic PRS profiles within our UK Biobank testing cohort.** Each graph presents the projected proportion of individuals diagnosed with a specific disease by a certain age, segmented into three PRS-based groups: the top 5%, representing high-risk individuals; the middle 25-75%, serving as the population reference; and the bottom 5%, indicating low-risk individuals.

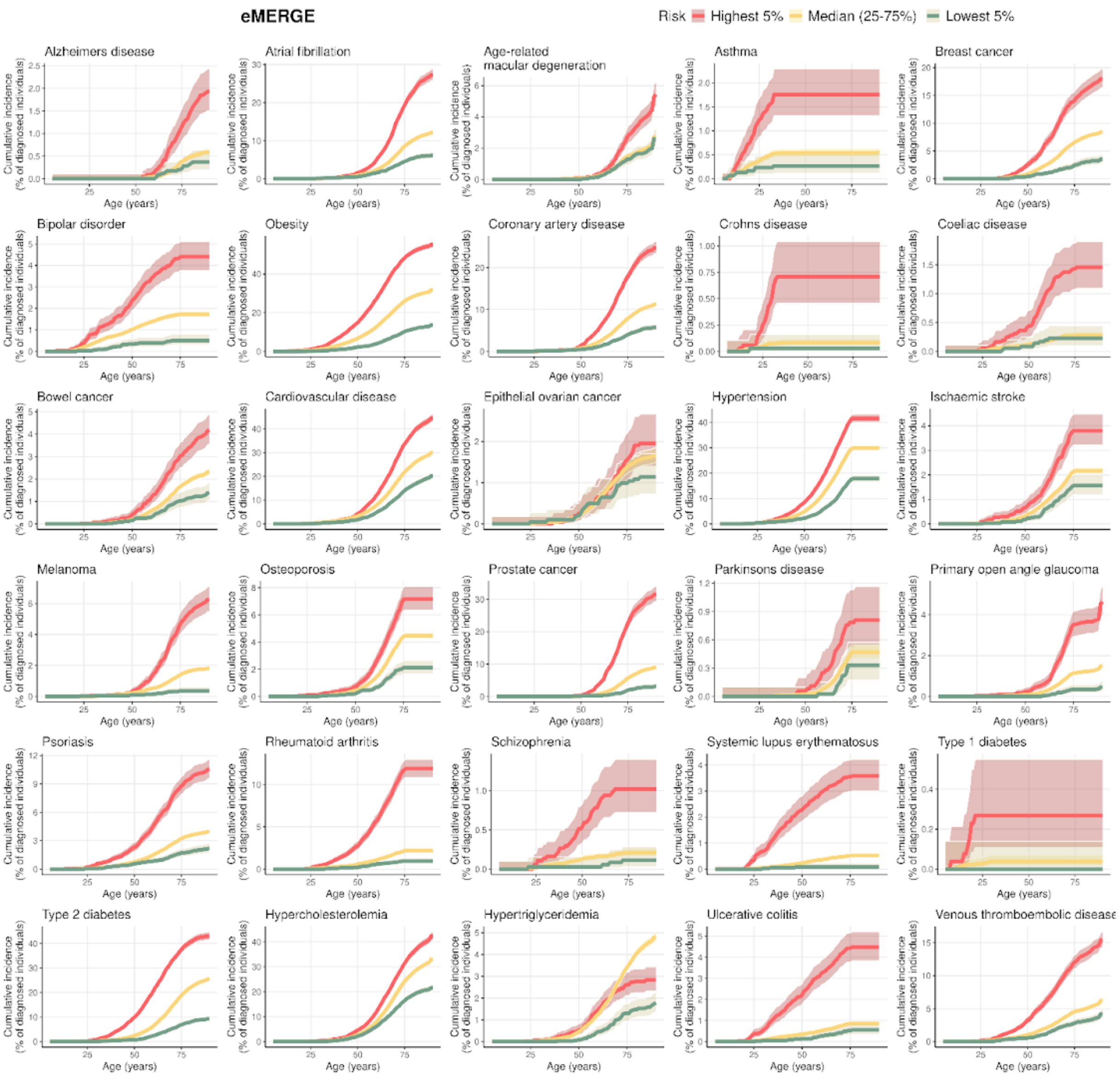
eMERGE
Risk
Highest 5%
Median (25-75%)
Lowest 5%
Alzheimers disease
Atrial fibrillation
Age-related macular degeneration
Asthma
Breast cancer
Bipolar disorder
Obesity
Coronary artery disease
Crohns disease
Coeliac disease
Bowel cancer
Cardiovascular disease
Epithelial ovarian cancer
Hypertension
Ischaemic stroke
Melanoma
Osteoporosis
Prostate cancer
Parkinsons disease
Primary open angle glaucoma
Psoriasis
Rheumatoid arthritis
Schizophrenia
Systemic lupus erythematosus
Type 1 diabetes
Type 2 diabetes
Hypercholesterolemia
Hypertriglyceridemia
Ulcerative colitis
Venous thromboembolic disease
Cumulative incidence (% of diagnosed individuals)
Age (years)

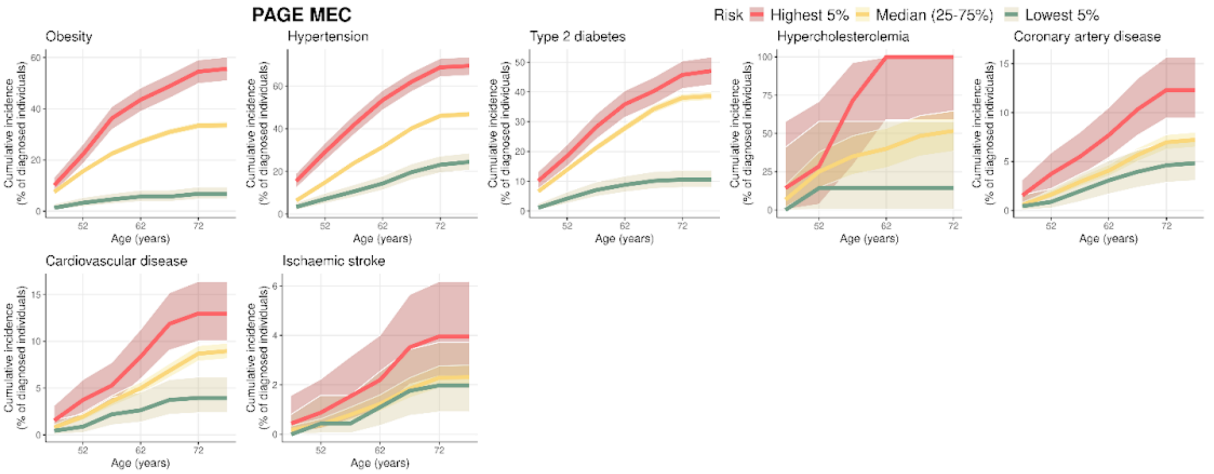
PAGE MEC
Risk
Highest 5%
Median (25-75%)
Lowest 5%
Obesity
Hypertension
Type 2 diabetes
Hypercholesterolemia
Coronary artery disease
Cardiovascular disease
Ischaemic stroke
Cumulative incidence (% of diagnosed individuals)
Age (years)

**Fig S8. Cumulative incidence graphs display the predictive accuracy of our polygenic PRS profiles within the eMERGE and PAGE MEC cohorts.** Each graph presents the projected proportion of individuals diagnosed with a specific disease by a certain age, segmented into three PRS-based groups: the top 5%, representing high-risk individuals; the middle 25-75%, serving as the population reference; and the bottom 5%, indicating low-risk individuals.

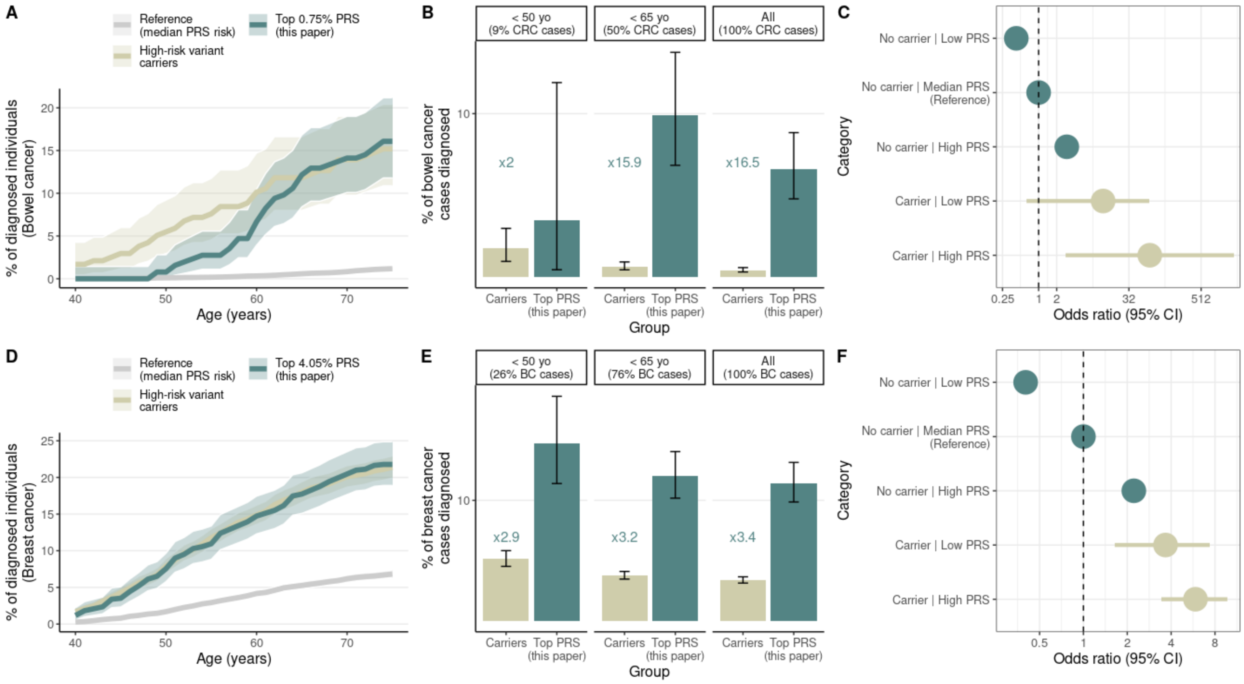


**Fig S9. Comparison between our PRS risk profiles and functional variant carriers in UKB Europeans.**

**A)** Cumulative incidence of colorectal cancer among carriers with pathogenic or likely-pathogenic mutations in cancer-related genes and contrasted it with individuals in the top 0.75% of our PRS distribution from the ensemble method, where a percentile chosen to match the risk up to age 70 with that of mutation carriers. Additionally, the median 25%-75% of the PRS distribution serves as the population reference. **B)** Percentage of colorectal cancer (CRC) cases diagnosed in individuals aged under 50, under 65, and across all age groups, occurring either in mutation carriers or in those at the top of the PRS distribution. The plots display the ratio of high PRS cases to mutation carrier cases in each age category. **C)** The forest plot displayed odds ratios for the observed risk in colorectal cancer, with horizontal lines representing the 95% confidence intervals. Non-carriers were categorized into three groups based on their polygenic scores—low, intermediate, and high, corresponding to the lowest 25%, middle 25-75%, and highest 25% of the distribution, respectively. The intermediate polygenic score group served as the reference category for calculating odds ratios. Due to the limited number of individuals with these mutations, carriers were only classified into lower and higher risk groups. **D)** Cumulative incidence of breast cancer among carriers with pathogenic or likely-pathogenic mutations in cancer-related genes and contrasted it with individuals in the top 4.05% of our PRS distribution from the ensemble method, where a percentile chosen to match the risk up to age 70 with that of mutation carriers. Additionally, the median 25%-75% of the PRS distribution serves as the population reference. **E)** Percentage of breast cancer (BC) cases diagnosed in individuals aged under 50, under 65, and across all age groups, occurring either in mutation carriers or in those at the top of the PRS distribution. The plots display the ratio of high PRS cases to mutation carrier cases in each age category. **F)** The forest plot displayed odds ratios for the observed risk in breast cancer, with horizontal lines representing the 95% confidence intervals. Non-carriers were categorized into three groups based on their polygenic scores—low, intermediate, and high, corresponding to the lowest 25%, middle 25-75%, and highest 25% of the distribution, respectively. The intermediate polygenic score group served as the reference category for calculating odds ratios. Due to the limited number of individuals with these mutations, carriers were only classified into lower and higher risk groups.

### Supplemental tables

**Table S1. Trait definition in UK Biobank and eMERGE cohorts.**

| | UK Biobank | | eMERGE |
|---|---|---|---|
| **Trait** | **Self-reported code and additional information** | **ICD-10 code** | **ICD-9 code** |
| **Age-related macular degeneration** | UKB code 1528 in field 20002; UKB code 5 in field 6148 | H35.3 | 362.50 |
| **Alzheimer's disease** | UKB code 1263 in field 20002 | F00, G30 | 331.0, 331.00 |
| **Asthma** | UKB code 1111 in field 20002 | J45, J46 | 493 |
| **Atrial fibrillation** | UKB code 1471 in field 20002 | I48 | 427.31 |
| **Bipolar disorder** | UKB code 1291 in field 20002 | F31 | 296.0, 296.1, 296.4, 296.5, 296.6, 296.7, 296.8 |
| **Bowel cancer** | UKB codes 1019, 1020, 1022, 1023 in field 20001 | C18, C19, C20 | 153, 154, V10.05, V10.06, 209.1 |
| **Breast cancer** | UKB code 1002 in field 20001 | C50 | V10.3, 174 |
| **Cardiovascular disease** | * UKB codes 1075, 1074, 1082, 1583 in field 20002<br>* codes 1,2,3 in field 6150<br>* UKB codes 1070, 1071, 1105, 1109, 1095 and 1514 in field 20004<br>* OPCS-4 codes K40-K46, K47.1, K49-K50, K75<br>* ICD9: 410-414, 434, 436 and 42979 | G45, I20-I25, I63-I64 | V12.54, 435, 437.7, 362.34, 410, 429.79, 429.71, 429.5, 429.6, 413, 411, 412, 414.1, 414.8, 414.9, 433.01, 433.11, 433.21, 433.31, 433.81, 433.91, 434.01, 434.11, 434.91 |
| **Celiac disease** | UKB code 1456 in field 20002 | K90.0 | 579.0 |
| **Coronary artery disease** | * UKB codes 1075 in field 20002<br>* code 1 in field 6150<br>* UKB codes 1070 and 1095 in field 20004<br>* OPCS-4 codes: K40.1–40.4, K41.1–41.4, K45.1–45.5, K49.1–49.2, K49.8–49.9, K50.2, K75.1–75.4, K75.8–75.9 | I21, I22, I23, I24.1, I25.2 | 410, 429.79, 429.71, 429.5, 429.6, 411.0, 412 |
| **Crohn's disease** | UKB code 1462 in field 20002 | K50, M07.4, M09.1 | 555 |
| **Epithelial ovarian cancer** | | C56 | 183.0, V10.43 |

| | | | |
|---|---|---|---|
| **Hypercholesterolemia** | UKB field 23400 (as quantitative trait: total cholesterol) | E78.0 (only eMERGE) | 272.0 |
| **Hypertension** | UKB codes 1065, 1072 in field 20002 | I10, I15 | 401, 405 |
| **Hypertrygliceridemia** | UKB field 23407 (as quantitative trait: triglyceride levels) | E78.1 (only eMERGE) | 272.1 |
| **Ischaemic stroke** | UKB code 1583 in field 20002 | I63, I64 | 433.01, 433.11, 433.21, 433.31, 433.81, 433.91, 434.01, 434.11, 434.91 |
| **Melanoma** | UKB code 1059 in field 20001 | C43 | 172 |
| **Obesity** | UKB field 21001 (as quantitative trait: body mass index) | E66 (only eMERGE) | 278.0, 649.1 |
| **Osteoporosis** | UKB code 1309 in field 20002 | M80, M81, M82 | 733.0 |
| **Prostate cancer** | UKB code 1044 in field 20001 | C61 | 185, V10.46 |
| **Parkinson's disease** | UKB code 1262 in field 20002 | G20 | 332.0 |
| **Primary open angle glaucoma** | UKB code 1277 in field 20002 | H40.1, H40.9 | 365.11, 365.9 |
| **Psoriasis** | UKB code 1453 in field 20002 | L40, L41 | 696.0, 696.1, 696.2, 696.8 |
| **Rheumatoid arthritis** | UKB code 1464 in field 20002 | M05, M06, M08.0 | 714.8, 714.9, 714.0, 714.2, 714.3, 714.1 |
| **Schizophrenia** | UKB code 1289 in field 20002 | F20 | 295, V11.0 |
| **Systemic lupus erythematosus** | UKB code 1381 in field 20002 | M32 | 710.0 |
| **Type 1 diabetes** | UKB code 1222 in 20002. See details in methods | E10 | 250.01, 250.03, 250.11, 250.13, 250.21, 250.23, 250.31, 250.33, 250.41, 250.43, 250.51, 250.53, 250.61, 250.63, 250.71, 250.73, 250.81, 250.83, 250.91, 250.93 |
| **Type 2 diabetes** | UKB code 1223 in 20002. See details in methods | E11 | 250.00, 250.02, 250.10, 250.12, 250.20, 250.22, 250.30, 250.32, 250.40, 250.42, 250.50, 250.52, 250.60, 250.62, 250.70, 250.72, 250.80, 250.82, 250.90, 250.92 |
| **Ulcerative colitis** | UKB code 1463 in field 20002 | K51, M07.5, M09.2 | 556 |
| **Venous thromboembolic disease** | UKB codes 1068, 1094, 1093 in field 20002 | I81, I82, I26, O22.3, O87.1, O08.2 | 452, 453, V12.51, 415.1, 671.42, 671.44, 639.6 |

**Table S2 | Studies and database sources for the GWAS meta-analysis for the 30 medically-related traits, including the numbers of cases and controls in each dataset.**

| Trait | Study/Database | Cases | Controls |
|---|---|---|---|
| **Age-related macular degeneration** | UKB[14] | 7,476 | 328,367 |
| | FinnGen[23] | 7,582 | 348,936 |
| | Guindo-Martínez *et al.* (2021)[27] | 3,685 | 52,952 |
| | Fritsche *et al.* (2016)[73] | 16,144 | 17,832 |
| | ∑ | **34,887** | **748,087** |
| **Alzheimer's disease** | UKB[14] | 1,760 | 335,351 |
| | FinnGen[23] | 13,393 | 363,884 |
| | Moreno-Grau *et al.* (2019)[28] | 6063 | 6305 |
| | Lambert *et al.* (2021)[29] | 17,008 | 37,154 |
| | ∑ | **38,224** | **742,694** |
| **Asthma** | UKB[14] | 48,278 | 288,833 |
| | FinnGen[23] | 42,163 | 202,399 |
| | Demenais *et al*. (2018)[33] | 5,186 | 7,660 |
| | Guindo-Martínez *et al.* (2021)[27] | 9,209 | 47,428 |
| | Sakaue *et al*. (2021)/BioBank Japan[24] | 13,015 | 162,933 |
| | Chang *et al*. (2022)[34] | 3,876 | 2,607 |
| | ∑ | **121,727** | **711,860** |
| **Atrial fibrillation** | UKB[14] | 22,180 | 285,774 |
| | FinnGen[23] | 45,766 | 191,924 |
| | Sakaue *et al*. (2021)/BioBank Japan[24] | 4,150 | 155,540 |
| | Low *et al.* (2017)[30] | 8,180 | 28,612 |
| | Christophersen *et al*. (2017)[31] | 18,398 | 91,536 |
| | Choi *et al*. (2018)[32] | 2,781 | 4,959 |
| | ∑ | **101,455** | **758,345** |
| **Bipolar disorder** | UKB[14] | 2,305 | 334,806 |
| | Mullins et al. (2021)[36] | 40,463 | 313,436 |
| | ∑ | **42,768** | **648,242** |
| **Bowel cancer** | UKB[14] | 6,171 | 302,196 |
| | FinnGen[23] | 5,458 | 287,137 |
| | Sakaue *et al*. (2021)/BioBank Japan[24] | 8,305 | 159,386 |
| | Tanikawa *et al*. (2018)[41] | 6,692 | 27,178 |
| | Garcia-Etxebarria *et al*. (2022)[40] | 835 | 940 |
| | Fernandez-Rozadilla *et al*. (2023)[42] | 100,204 | 154,587 |
| | ∑ | **127,665** | **931,424** |
| **Breast cancer** | UKB[14] | 12,933 | 152,004 |
| | FinnGen[23] | 15,680 | 167,189 |

| Trait | Study/Database | | Cases | Controls |
|---|---|---|---|---|
| | Sakaue *et al.* (2021)/BioBank Japan[24] | | 6,325 | 73,225 |
| | Michailidou *et al.* (2015)[35] | | 46,785 | 42,892 |
| | | ∑ | **81,723** | **435,310** |
| **Cardiovascular disease** | UKB[14] | | 37,890 | 269,427 |
| | FinnGen[23] | | 185,353 | 191,924 |
| | Kanai *et al.* (2021)/BioBank Japan[25] | | 32,512 | 146,214 |
| | Guindo-Martínez *et al.* (2021)[27] | | 15,009 | 41,628 |
| | Nikpay *et al.* (2015)[29] | | 60,801 | 123,504 |
| | | ∑ | **331,565** | **772,697** |
| **Celiac disease** | UKB[14] | | 2,656 | 334,455 |
| | FinnGen[23] | | 3,690 | 361,055 |
| | | ∑ | **6,346** | **695,510** |
| **Coronary artery disease** | UKB[14] | | 31,451 | 276,086 |
| | FinnGen[23] | | 47,550 | 313,400 |
| | Kanai *et al.* (2021)/BioBank Japan[25] | | 32,512 | 146,214 |
| | Nikpay *et al.* (2015)[37] | | 60,801 | 123,504 |
| | | ∑ | **172,314** | **859,204** |
| **Crohn's disease** | UKB[14] | | 2,064 | 335,047 |
| | FinnGen[23] | | 1,665 | 375,445 |
| | de Lange *et al.* (2017)[38] | | 4,474 | 9,500 |
| | Garcia-Etxebarria *et al.* (2022)[39] | | 284 | 935 |
| | | ∑ | **8,487** | **720,927** |
| **Epithelial ovarian cancer** | UKB[14] | | 1,226 | 179,806 |
| | Sakaue *et al.* (2021)/BioBank Japan[24] | | 843 | 60,614 |
| | Phelan *et al.* (2017) [43] | | 16,924 | 68,502 |
| | Lawrenson *et al.* (2019) [44] | | 3,238 | 4,083 |
| | | ∑ | **22,231** | **313,005** |
| **Hypercholesterolemia (Total cholesterol)** | UKB[14] | | 83,838 | |
| | Graham *et al.* (2021)[70] | | 1,251,910 | |
| | | ∑ | **1,335,748** | |
| **Hypertension** | UKB[14] | | 121,548 | 184,542 |
| | FinnGen[23] | | 111,581 | 265,626 |
| | Guindo-Martínez *et al.* (2021)[27] | | 28,391 | 28,246 |
| | Wojcik *et al.* (2019)[26] | | 27,123 | 22,018 |
| | Oh *et al.* (2020)[45] | | 1,864 | 7,812 |
| | | ∑ | **290,507** | **508,244** |
| **Hypertriglyceridemia (Total triglycerides)** | UKB[14] | | 83,838 | |
| | Graham *et al.* (2021)[70] | | 1,121,280 | |

| Trait | Study/Database | Cases | Controls |
|---|---|---|---|
| | | ∑ **1,205,118** | |
| **Ischemic stroke** | UKB[14] | 5,761 | 302,890 |
| | FinnGen[23] | 39,818 | 271,817 |
| | Malik *et al.* (2018) * only EUR[46] | 34,217 | 406,111 |
| | Mishra *et al.* (2022) * only non-EUR[47] | 24,568 | 269,090 |
| | | ∑ **104,364** | **1,249,908** |
| **Melanoma** | UKB[14] | 4,168 | 331,681 |
| | FinnGen[23] | 3,960 | 286,874 |
| | Sakaue *et al.* (2021)/BioBank Japan[24] | 154 | 178,572 |
| | Landi *et al.* (2020)[48] | 30,143 | 81,405 |
| | | ∑ **38,425** | **878,532** |
| **Obesity (BMI)** | UKB[14] | 336,073 | |
| | FinnGen[23] | 412,181 | |
| | Sakaue *et al.* (2021)/BioBank Japan[24] | 163,835 | |
| | Wojcik *et al.* (2019)[26] | 49,335 | |
| | | ∑ **961,424** | |
| **Osteoporosis** | UKB[14] | 14,636 | 321,851 |
| | FinnGen[23] | 7,300 | 358,014 |
| | Sakaue *et al.* (2021)/BioBank Japan[24] | 9,794 | 168,932 |
| | Guindo-Martínez *et al.* (2021)[27] | 5,399 | 51,238 |
| | | ∑ **37,129** | **900,035** |
| **Parkinson's disease** | UKB[14] | 2,343 | 334,768 |
| | FinnGen[23] | 4,235 | 373,042 |
| | Sakaue *et al.* (2021)/BioBank Japan[24] | 340 | 175,788 |
| | Nalls *et al.* (2019)[51] | 15,056 | 12,637 |
| | Rodrigo *et al.* (2021)[52] | 5,167 | 5,366 |
| | Le Guen *et al.* (2021)[53] | 3,500 | 299,408 |
| | | ∑ **30,641** | **1,201,009** |
| **Primary open angle glaucoma** | UKB[14] | 7,674 | 326,897 |
| | FinnGen[23] | 7,756 | 358,375 |
| | Sakaue *et al.* (2021)/BioBank Japan[24] | 8,448 | 168,903 |
| | Gharahkhani *et al.* (2021)[54] | 6,935 | 39,588 |
| | | ∑ **30,813** | **893,763** |
| **Prostate cancer** | UKB[14] | 9,687 | 140,038 |
| | FinnGen[23] | 13,216 | 119,948 |
| | Sakaue *et al.* (2021)/BioBank Japan[24] | 5,672 | 84,660 |
| | Schumacher *et al.* (2018)[49] | 79,148 | 61,106 |
| | Ito *et al.* (2023)[50] | 107,281 | 197,733 |
| | | ∑ **215,004** | **603,485** |

| Trait | Study/Database | Cases | Controls |
|---|---|---|---|
| **Psoriasis** | UKB[14] | 6,358 | 330,753 |
| | FinnGen[23] | 9,267 | 364,071 |
| | Sakaue *et al.* (2021)/BioBank Japan[24] | 206 | 172,289 |
| | Stuart *et al.* (2021)[55] | 18,557 | 29,914 |
| | ∑ | **34,388** | **897,027** |
| **Rheumatoid arthritis** | UKB[14] | 7,719 | 329,392 |
| | FinnGen[23] | 12,555 | 240,862 |
| | Ishigaki *et al.* (2022)[56] | 35,871 | 1,698,689 |
| | ∑ | **56,145** | **2,268,943** |
| **Schizophrenia** | UKB[14] | 698 | 336,413 |
| | FinnGen[23] | 6,515 | 364,160 |
| | Sakaue *et al.* (2021)/BioBank Japan[24] | 99 | 177,794 |
| | Trubetskoy *et al.* (2022)[60] | 67,323 | 93,456 |
| | ∑ | **74,635** | **971,823** |
| **Systemic lupus erythematosus** | UKB[14] | 584 | 336,527 |
| | FinnGen[23] | 652 | 353,088 |
| | Sakaue *et al.* (2021)/BioBank Japan[24] | 317 | 175,937 |
| | Langefeld *et al.* (2017)[57] | 6,748 | 11,516 |
| | Wang *et al.* (2021)[58] | 8,798 | 16,470 |
| | Song *et al.* (2021)[59] | 512 | 994 |
| | ∑ | **17,611** | **894,532** |
| **Type 1 diabetes** | UKB[14] | 1,200 | 333,542 |
| | FinnGen[23] | 8,967 | 308,373 |
| | Sakaue *et al.* (2021)/BioBank Japan[24] | 1,219 | 132,032 |
| | Inshaw *et al.* (2021)[61] | 7,467 | 10,218 |
| | Robertson *et al.* (2021)[62] | 22,153 | 37,374 |
| | ∑ | **41,006** | **821,539** |
| **Type 2 diabetes** | UKB[14] | 24,800 | 281,093 |
| | FinnGen[23] | 57,698 | 308,252 |
| | Chen *et al.* (2019)[63] | 2,633 | 1,714 |
| | Guindo-Martínez *et al.* (2021)[27] | 6,967 | 49,670 |
| | Mansour *et al.* (2021)[64] | 9,486 | 2,744 |
| | Spracklen *et al.* (2020)[65] | 77,418 | 356,122 |
| | Wojcik *et al.* (2019)[26] | 14,042 | 31,683 |
| | Million Veterans Program[66] | 102,683 | 170,726 |
| | Cai *et al.* (2020)[67] | 9,978 | 12,348 |
| | Loh *et al.* (2022)[68] | 16,677 | 33,856 |
| | Scott *et al.* (2017)[69] | 26,676 | 132,532 |
| | ∑ | **349,058** | **1,380,740** |

| Trait | Study/Database | | Cases | Controls |
|---|---|---|---|---|
| **Ulcerative colitis** | UKB[14] | | 3,914 | 333,197 |
| | FinnGen[23] | | 5,034 | 371,530 |
| | Liu *et al.* (2015)[71] | | 6,968 | 20,464 |
| | | ∑ | **15,916** | **725,191** |
| **Venous thromboembolism** | UKB[14] | | 13,934 | 323,177 |
| | FinnGen[23] | | 19,372 | 357,905 |
| | Million Veterans Program[66] | | 11,844 | 211,753 |
| | Ghouse *et al.* (2023)[72] | | 81,190 | 1,419,671 |
| | | ∑ | **126,340** | **2,312,506** |

**Table S3 | Risk factors included in the final PRS-based disease prediction model.**

| Trait | Additional Risk Factors | | | | | | | | | | | |
|---|---|---|---|---|---|---|---|---|---|---|---|---|
| | BMI | BC | CAD | CD | CED | CVD | HT | ISS | RA | T1D | T2D | UC |
| Age-related macular degeneration | ✓ | / | / | / | / | / | ✓ | / | / | / | / | / |
| Alzheimer's disease | ✓ | / | / | / | / | / | ✓ | ✓ | / | ✓ | ✓ | / |
| Asthma | ✓ | / | / | / | / | / | / | / | / | / | / | / |
| Atrial fibrillation | ✓ | / | ✓ | / | / | ✓ | ✓ | / | / | / | ✓ | / |
| Bipolar disease | / | / | / | / | / | / | / | / | / | / | / | / |
| Bowel cancer | ✓ | / | / | ✓ | / | / | / | / | / | ✓ | ✓ | ✓ |
| Breast cancer | ✓ | / | / | / | / | / | / | / | / | / | / | / |
| Cardiovascular disease | ✓ | / | / | / | / | / | ✓ | / | / | ✓ | ✓ | / |
| Celiac disease | / | / | / | / | / | / | / | / | / | ✓ | / | / |
| Coronary artery disease | ✓ | / | / | / | / | / | ✓ | / | / | ✓ | ✓ | / |
| Crohn's disease | / | / | / | / | / | / | / | / | / | / | / | / |
| Epithelial ovarian cancer | ✓ | ✓ | / | / | / | / | / | / | / | ✓ | / | ✓ |
| Hypercholesterolemia | ✓ | / | / | / | / | / | / | / | / | / | / | / |
| Hypertension | ✓ | / | / | / | / | / | / | / | / | ✓ | ✓ | / |
| Hypertrygliceridemia | ✓ | / | / | / | / | / | / | / | / | / | / | / |
| Ischemic stroke | ✓ | / | / | / | / | / | ✓ | / | / | / | / | / |
| Melanoma | / | / | / | / | / | / | / | / | / | / | / | / |
| Obesity | / | / | / | / | / | / | / | / | / | / | / | / |
| Osteoporosis | / | / | / | ✓ | ✓ | / | / | / | ✓ | / | / | ✓ |
| Parkinson's disease | / | / | / | / | / | / | / | / | / | / | / | / |
| Primary open angle glaucoma | / | / | / | / | / | / | ✓ | / | / | ✓ | ✓ | / |
| Prostate cancer | ✓ | / | / | / | / | / | / | / | / | / | / | / |
| Psoriasis | ✓ | / | / | / | / | / | ✓ | / | / | / | / | / |
| Rheumatoid arthritis | ✓ | / | / | / | / | / | / | / | / | / | / | / |
| Schizophrenia | / | / | / | / | / | / | / | / | / | / | / | / |
| Systemic lupus erythematosus | / | / | / | / | / | / | / | / | / | / | / | / |
| Type 1 diabetes | / | / | / | / | / | / | / | / | / | / | / | / |
| Type 2 diabetes | ✓ | / | / | / | / | / | / | / | / | / | / | / |
| Ulcerative colitis | / | / | / | / | / | / | / | / | / | / | / | / |
| Venous thromboembolism | ✓ | / | ✓ | / | / | ✓ | ✓ | / | / | ✓ | ✓ | / |

BMI = body mass index; CAD = Coronary artery disease; CD = Crohn's disease; CED = Celiac disease; CVD = Cardiovascular disease; HT = Hypertension; ISS = Ischemic stroke; RA = Rheumatoid arthritis; T1D = Type 1 diabetes; T2D = Type 2 diabetes; UC = Ulcerative colitis.
